# CalScope: Monitoring SARS-CoV-2 Seroprevalence from Vaccination and Prior Infection in Adults and Children in California May 2021– July 2021

**DOI:** 10.1101/2021.12.09.21267565

**Authors:** Megha L. Mehrotra, Esther Lim, Katherine Lamba, Amanda Kamali, Kristina W. Lai, Erika Meza, Irvin Szeto, Peter Robinson, Cheng-ting Tsai, David Gebhart, Noemi Fonseca, Andrew B. Martin, Catherine Ley, Steve Scherf, James Watt, David Seftel, Julie Parsonnet, Seema Jain

## Abstract

**Importance:** Understanding how SARS-CoV-2 seroprevalence varies regionally across California is critical to the public health response to the pandemic.

**Objective:** To estimate how many Californians have antibodies against SARS-CoV-2 from prior infection or vaccination.

**Design:** Wave 1 of CalScope: a repeated cross-sectional serosurvey of adults and children enrolled between April 20, 2021 and June 16, 2021.

**Setting:** A population-based random sample of households in seven counties in California (Alameda, El Dorado, Kern, Los Angeles, Monterey, San Diego, and Shasta) were invited to complete an at-home SARS-CoV-2 antibody test and survey instrument.

**Participants:** Invitations were sent to 200,000 randomly selected households in the seven counties. From each household, 1 adult (18 years and older) and 1 child (aged 6 months to 17 years) could enroll in the study. There were no exclusion criteria.

**Main Outcome(s) and Measures:** All specimens were tested for antibodies against the nucleocapsid and spike proteins of SARS-CoV-2. The primary outcome was serostatus category, which was determined based on antibody test results and self-reported vaccination status: seronegative, antibodies from infection only, antibodies from infection and vaccination, and antibodies from vaccination alone. We used inverse probability of selection weights and iterative proportional fitting to account for non-response.

**Results:** 11,161 households enrolled in wave 1 of CalScope, with 7,483 adults and 1,375 children completing antibody testing. As of June 2021, 27% (95%CI [23%, 31%]) of adults and 30% (95%CI [24%, 36%]) of children had evidence of prior SARS-CoV-2 infection; 33% (95%CI [28%, 37%]) of adults and 57% (95%CI [48%, 66%]) of children were seronegative. Serostatus varied regionally. Californians 65 years or older were most likely to have antibodies from vaccine alone (59%; 95%CI [48%, 69%]) and children between 5-11 years old were most likely to have antibodies from prior infection alone (36%; 95%CI [21%, 52%]).

**Conclusions and Relevance:** As of June 2021, a third of adults in California and most children under 18 remained seronegative. Seroprevalence varied regionally and by demographic group, suggesting that some regions or populations might remain more vulnerable to subsequent surges than others.

**Key Points:** *Question:* What is the prevalence of vaccine and infection derived antibodies against SARS-CoV-2 in adults and children in California?

*Findings:* In this population-based serosurvey that included 11,161 households, as of June 2021, 33% of adults and 57% of children were seronegative; 18% of adults and 26% of children had antibodies from infection alone; 9% of adults and 5% of children had antibodies from both infection and vaccination; and 41% of adults and 13% of children had antibodies from vaccination alone.

*Meaning:* Serostatus varied considerably across geographic regions, suggesting that certain areas might be at increased risk for future COVID-19 surges.

## INTRODUCTION

By July 2021, the United States had recorded more than 34 million COVID-19 cases and 600,000 deaths, with over 3.7 million cases and 60,000 deaths in California.^1^ Though all adults and children over 12 have been eligible for COVID-19 vaccination since May 2021 in California, vaccine uptake has been uneven; as of July 31, 2021, the percent of persons fully vaccinated ranged from 24% to 79% across California counties.

The California Department of Public Health (CDPH) monitors COVID-19 burden and forecasts hospitalizations to determine when additional mitigation measures are required to avoid overwhelming the healthcare system.^2^ Both prior SARS-CoV-2 infection and vaccination reduce the risk of symptomatic COVID-19 and hospitalization, although questions remain regarding the relative level and duration of risk reduction.^3–5^ Accurately forecasting future COVID-19 surges requires estimating population immunity from prior infection or vaccination in order to determine how many people remain susceptible to infection. Estimating population immunity using routine surveillance data is challenging. Since COVID-19 may be asymptomatic and persons with mild illness may not seek testing, many infections are not recognized or reported. Recent studies estimated that 70% of SARS-CoV-2 infections in California were unaccounted for in the CDPH COVID-19 surveillance system by December 2020.^6^

Population-based serosurveys can estimate immunity from prior infection or vaccination without the limitations inherent in routine surveillance, and several seroprevalence studies have been completed or are currently underway throughout the United States.^7–13^ However, the studies conducted thus far in California have been limited to convenience samples, restricted to narrow geographic regions, or only powered to produce statewide estimates, thereby limiting their utility for informing public health policy regionally in California.^6, 14, 15^ Thus, CDPH launched a population-based serosurvey (CalScope) to estimate the proportion of housed and non-institutionalized Californians with evidence of immunity against SARS-CoV-2 from prior infection or vaccination.

## METHODS

### Study Design

CalScope is a repeated cross-sectional study using random address-based sampling of households in seven counties in California. The study re-samples households with replacement over three timepoints.

### Sampling Strategy

We used a multistage sampling strategy to allow for region-specific seroprevalence estimates. The sampling approach was guided by principles of causal transportability^16^ to ensure that the final study results could be appropriately and efficiently generalized to the general population (Appendix). We sampled households in seven counties: Alameda, El Dorado, Kern, Los Angeles, Monterey, San Diego, and Shasta.

We used an address-based sampling frame created by Marketing Systems Group to select a probability sample of households within each county. The frame uses the United States Postal Service Computerized Delivery Sequence File, which covers all residential delivery locations in the United States, with each address geocoded and linked to the 2015 American Community Survey.^17^ We oversampled households from census tracts with higher proportions of Black households to ensure adequate representation. To enroll a total of 10,000 households, we sampled 200,000 households per wave distributed across the seven counties proportional to each county’s population with a minimum of 15,000 households sampled per county.

Sampled households could enroll one adult and one child (6 months to 17 years old). To randomize which eligible household members participated, we instructed households to enroll the adult and child with the next upcoming birthday. Wave 1 enrollment was conducted from April 20, 2021 through June 15, 2021.

### Survey Instruments

When registering for the study, participants completed a household enumeration form and elected to order at-home antibody test kits. Participants who declined the antibody test could choose to only complete the survey instrument.

The adult survey asked about demographics of all household members, income, occupation, medical history, COVID-19 vaccination and testing history, and behaviors associated with COVID-19 risk—including mask use and social distancing. The child survey asked about the child participant’s demographics, medical and COVID-19 disease history, and attendance in school and other social activities. (Appendix)

### Antibody Testing

Participants were mailed at-home antibody test kits with instructions on how to collect a dried blood spot (DBS) specimen and were asked to return their sample to Enable Biosciences within 30 days. Specimens with inadequate volume or collected >30 days before receipt by the laboratory were rejected. All valid specimens received by the laboratory by August 1, 2021 were included in this analysis.

Specimens were tested for both anti-spike and anti-nucleocapsid antibodies using Enable’s ADAP SARS-CoV-2 total antibody assay. The assay procedures have been described previously (Appendix).^18^ The assay cutoffs were established by testing 100 healthy controls and set at 99.7% percentile. The cutoffs for spike and nucleocapsid antibodies were 3.00 ΔCt and 1.50 ΔCt respectively. The assay was previously shown to be 100% sensitive and 100% specific in validation studies using DBS samples against the spike protein and nucleocapsid proteins.^18^

### Sampling Weights

We anticipated that households that enrolled in the study and completed antibody tests would differ from those that did not respond. Thus, we constructed sampling weights to generalize our results from the study sample to the target population: the general population of non-institutionalized, housed residents in each of the seven sampled counties.^19,20^

Within each county, there were three levels of selection between the final study sample and target population (Figure 1). The sampled population was all households that were mailed invitations to participate in CalScope; the registered population included all participants that registered for the study and completed a survey instrument; and the final study sample included all registered participants who completed an antibody test. We estimated weights to generalize across each selection step: Step 1) from the final study sample to the registered population, Step 2) from the registered population to the sampled population, and Step 3) from the sampled population to the target population.

**Figure 1.**
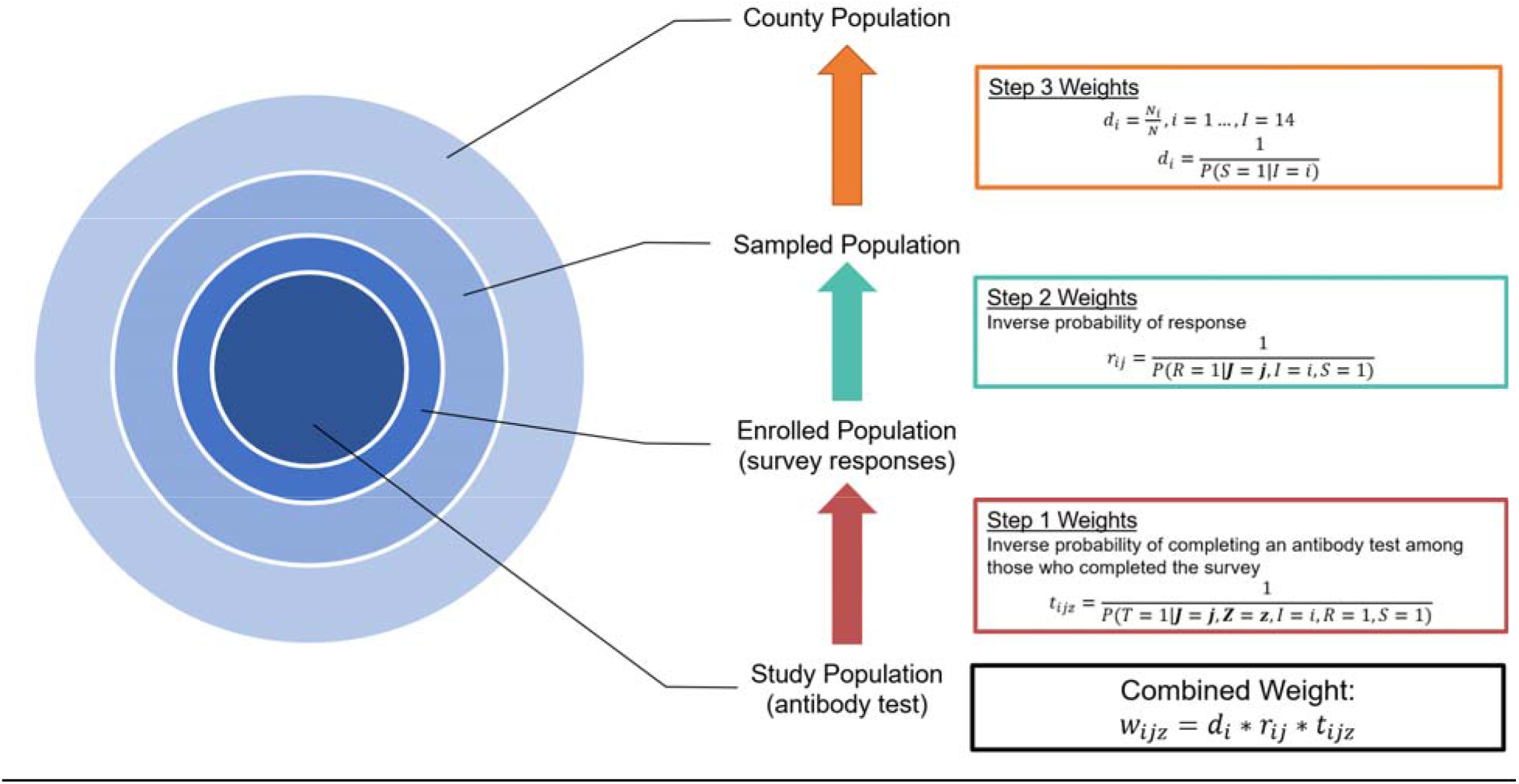
Levels of selection between study sample and target population in each county and corresponding weighting steps. In the <u>Step 3 Weights</u>, S=1 indicates that the ousehold was sampled and I=i is the sampling stratum. In the *Step 2 Weights*, R=1 indicates that at least one member of the household completed a survey instrument (enrolled). **J** is a vector of address-based characteristics including demographic characteristics from the American Community Survey, Healthy Places Index quartile, 2020 Presidential Election results by voter precinct, and COVID-19 vaccination coverage as of April 20, 2021 by zip code. In the <u>Step 1 Weights</u>, T=1 indicates that an individual has a valid antibody test result, **Z** is a vector of individual-level measurements from the survey instrument. Step 3 and 2 weights were estimated at the household level. Step 1 weights were estimated at the individual level with weights estimated separately for adults and children. The combined weight is the product of all 3 weights.

We constructed selection diagrams to guide variable selection for estimating the weights in steps 1 and 2 (Supplementary Figure 1).^16, 21^ Candidate variables for Step 1 included items from the survey instrument including: participant demographics, SARS-CoV-2 testing and vaccination history, mask use, ability to work remotely, household income, education, whether anyone in the household was considered an essential worker^22^, and any known contacts with a COVID-19 case. Candidate variables also included neighborhood-level characteristics from the 2015 American Community Survey (poverty, crowded living conditions, income, education, and race/ethnicity),^17^ ZIP-code level COVID-19 vaccination coverage as of May 2021, and 2020 Presidential general election results by voting precinct.^23^ Finally, we included the Healthy Places Index (HPI), a summary measure of neighborhood conditions that are associated with life-expectancy.^24^ Residents in neighborhoods in HPI quartile 4 have shorter life expectancies compared to those in HPI quartiles 1 to 3. Because we did not have survey responses from sampled households that never registered for the study, candidate variables for the Step 2 weights were limited to the neighborhood-level characteristics listed above.

We used a cross-validated ensemble machine learning algorithm, SuperLearner,^25^ to estimate inverse probability of selection weights for both Step 1 and Step 2. We included a mixture of parametric and machine learning algorithms in the SuperLearner. Weights for Step 1 were estimated separately for adults and children within each county. Step 2 weights were estimated at the household level within each county. Finally, we used the known sampling probabilities for each invited household to construct the Step 3 weights.

We multiplied all three weights and used iterative proportional fitting (raking) to calibrate the combined weights to ensure that the weighted distribution of age, sex, race/ethnicity, education, household income, and COVID-19 vaccination coverage matched the marginal distributions in the 2015 ACS and the state COVID-19 vaccine registry in each county.^26^

### Primary Outcomes

Participation in CalScope was anonymous, so we could not verify participants’ vaccination status. Instead, we used self-reported vaccination status, anti-nucleocapsid, and anti-spike antibody results, to classify participants into 4 mutually exclusive serostatus categories: <u>1) Seronegative</u>: negative nucleocapsid test and negative spike test regardless of self-reported vaccination status; <u>2) Prior Infection Only</u>: positive nucleocapsid test AND negative spike test OR (positive nucleocapsid test OR positive spike test) AND self-reported not having received any doses of a COVID-19 vaccine; <u>3) Infected and Vaccinated</u>: Positive nucleocapsid test AND positive spike tests AND self-reported at least 1 dose of any COVID-19 vaccine; <u>and 4) Vaccinated only:</u> Negative nucleocapsid test AND positive spike test AND self-reported at least 1 dose of any COVID-19 vaccine.

Using the sampling weights, we estimated the proportion of the population in each serostatus category and with evidence of prior infection for the whole sample and stratified by county, age, race/ethnicity, and HPI quartile. We used a non-parametric bootstrap with 1000 replicates to obtain 95% confidence intervals.

We estimated the ratio of the number of SARS-CoV-2 infections to confirmed cases in the CDPH’s COVID-19 case registry in the overall sample and stratified by county for both adults and children. To do this, we divided the proportion of the population with evidence of prior infection in CalScope by the proportion of the population that was a confirmed COVID-19 case as of 14 days prior to the median specimen collection date. A confirmed COVID-19 case was defined as a person with a positive PCR SARS-CoV-2 test; the cutoff date allowed for approximately 14 days between time of infection to seroconversion.

All analyses were conducted in R version 3.6.0 using the sl3 package for SuperLearner implementation, the anesrake package for iterative proportional fitting, and the survey package for analysis of the weighted data.^25, 27, 28^

## RESULTS

Of the 200,000 households invited, 11,161 registered for the study (5.6%) (Figure 2). 8,322 (74.6%) households completed an adult survey and 7,751 households (69%) completed adult antibody testing. 7,483 (67%) adults completed the survey and returned a DBS specimen with valid antibody results. Of the 11,161 households that registered for the study, 3,388 (30%) included at least one eligible child. A total of 2,013 child surveys (65%) and 1,436 (42.4%) child antibody tests were completed, and 1,375 (40.6%) children completed both the survey and an antibody test. Table 1 shows the demographics of the study sample before and after weighting. The median specimen collection date was May 22, 2021, with 60% of specimens collected in May 2021 and 90% of specimens collected in May or June 2021. (Supplementary Figure 2)

**Figure 2.**
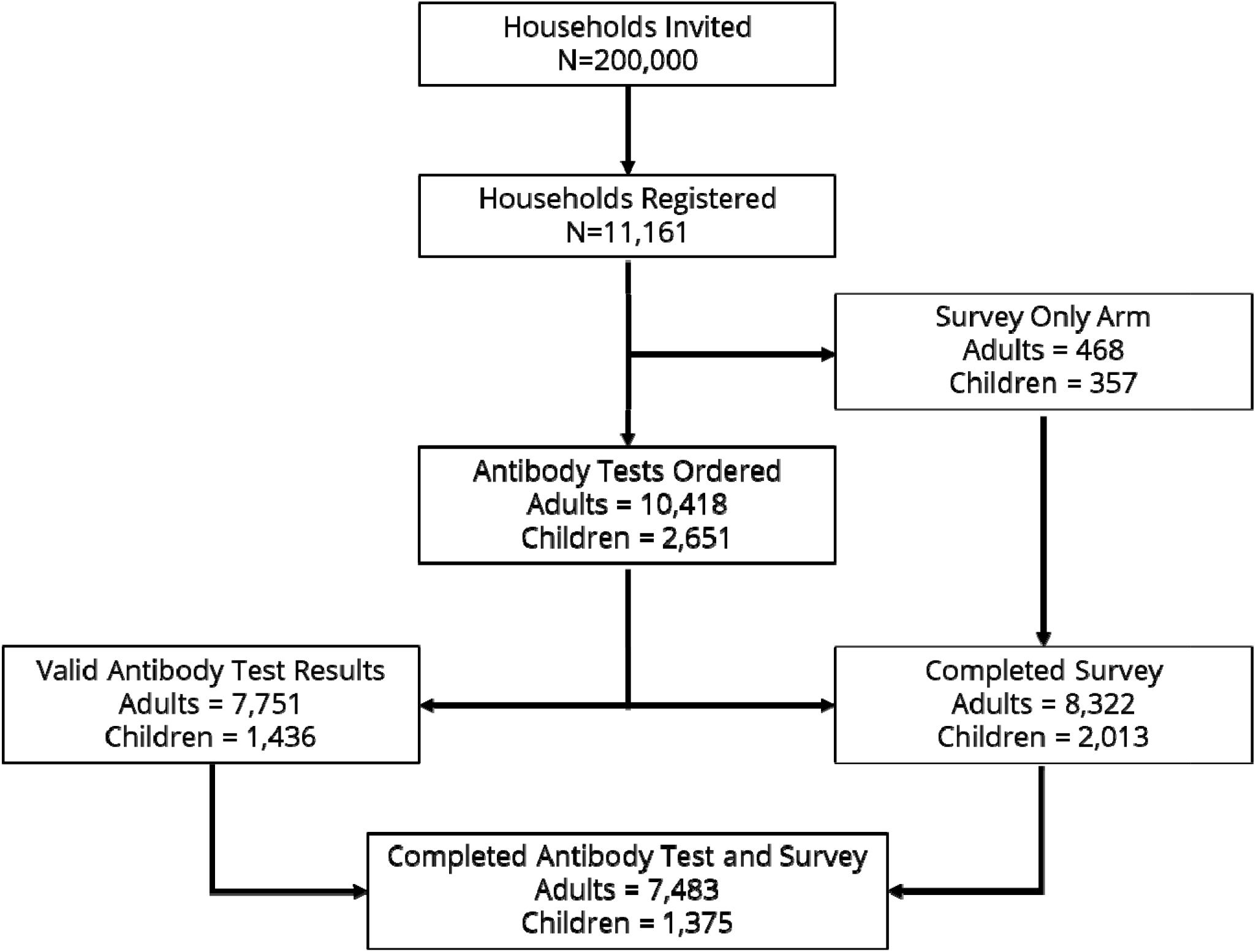
Wave 1 Consort Diagram. The final study sample includes those who completed an antibody test and survey instrument.

**TABLE 1.**
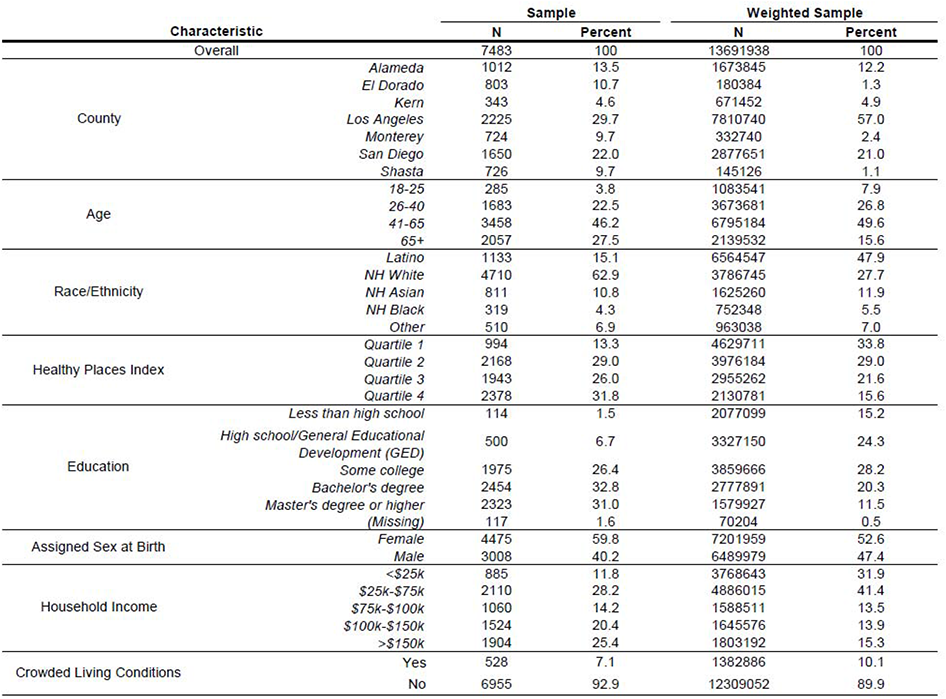
Study Sample

### Spike and Nucleocapsid Seroprevalence

Overall, 6,625/7,483 (89%) adults and 581/1,375 (42%) children had detectable spike antibodies; 846/7,483 (11%) adults and 224/1,375 (16%) children had detectable nucleocapsid antibodies. The weighted spike seroprevalence was 67% (95% CI [63%,71%]) for adults and 41% (95% CI [35%, 47%]) for children; the weighted nucleocapsid seroprevalence was 22% (95% CI [18%, 26%]) among adults and 25% (95% CI [19%, 31%]) in children. (Table 2)

**TABLE 2.**
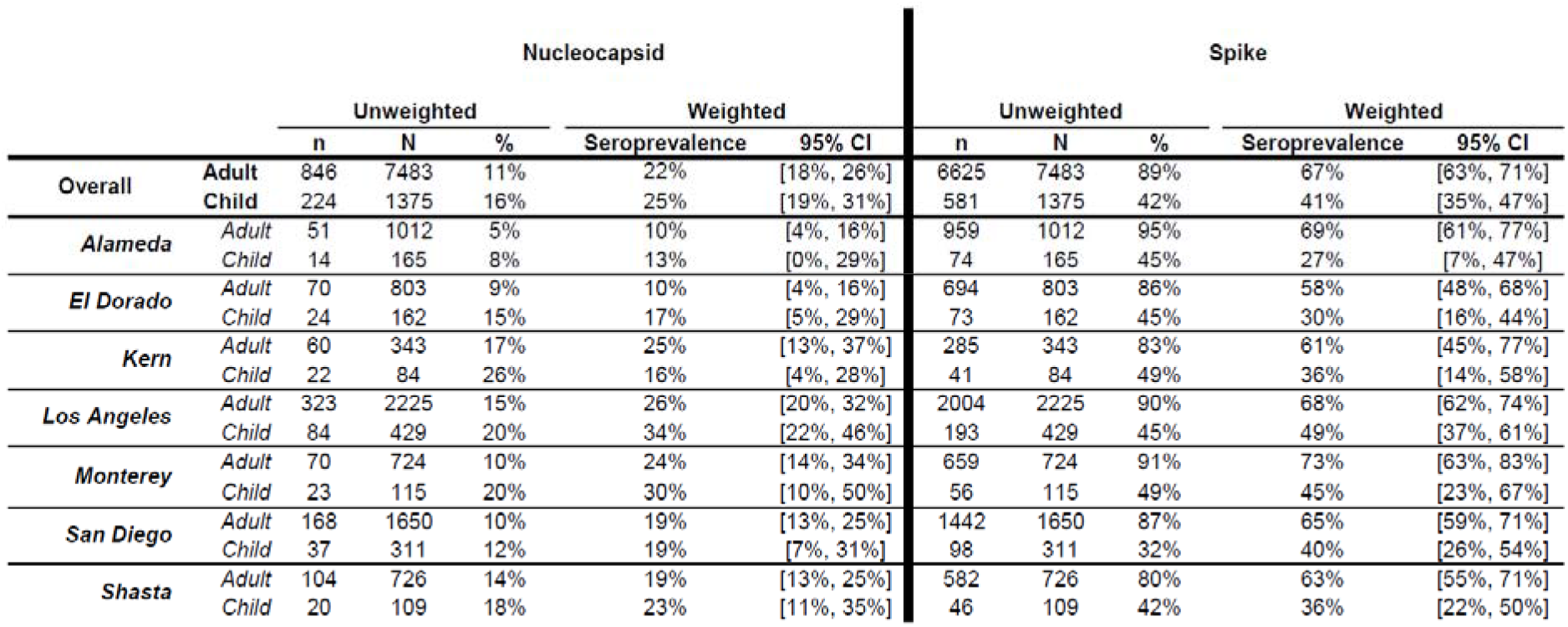
Nucleocapsid and Spike Results

### Serostatus

Among adults, we estimated that 33% (95% CI [28%, 37%]) were seronegative; 18% (95% CI [14%, 22%]) had antibodies from previous infection but not vaccination; 9% (95% CI [6%, 11%]) had antibodies from prior infection and vaccination; and 41% (95% CI [37%, 45%]) had antibodies from vaccination alone. (Table 3) Among children, 57% (95% CI [48%, 67%]) were seronegative; 26% (95% CI [19%, 32%]) had antibodies from prior infection but not vaccination; 5% (95% CI [1%, 8%]) had antibodies from prior infection and vaccination; and 13% (95% CI [7%, 18%]) had antibodies from vaccination alone. (Table 4) There was considerable regional heterogeneity in serostatus for adults and children across the seven counties. For example, seronegativity in adults varied from 27% (95% CI [15%, 38%]) in Monterey county to 42% (95% CI [29%, 54%]) in El Dorado county. For children, seronegativity ranged from 51% (95% CI [34%, 67%]) in Los Angeles County to 68% (95% CI [41%, 96%]) in El Dorado County.

**TABLE 3.** ADULT SEROSTATUS BY REGION, AGE, RACE/ETHNICITY. AND HPI QUARTILE

| Characteristic |  | Evidence of Prior Infection |  | Serostatus |  |  |  |  |  |  |  |
| --- | --- | --- | --- | --- | --- | --- | --- | --- | --- | --- | --- |
|  |  |  |  | Seronegative |  | Prior Infection Only |  | Prior Infection and Vaccination |  | Vaccination Only |  |
|  |  | Weighted Percent | [95% CI] | Weighted Percent | [95% CI] | Weighted Percent | [95% CI] | Weighted Percent | [95% CI] | Weighted Percent | [95% CI] |
| Overall |  | 27% | [23%, 31%] | 33% | [28%, 37%] | 18% | [14%, 22%] | 9% | [6%, 11%] | 41% | [37%, 45%] |
| County | Alameda | 19% | [11%, 27%] | 31% | [20%, 41%] | 15% | [6%, 23%] | 4% | [1%, 7%] | 51% | [38%, 63%] |
|  | El Dorado | 14% | [8%, 20%] | 42% | [29%, 54%] | 10% | [4%, 16%] | 5% | [2%, 7%] | 44% | [34%, 54%] |
|  | Kern | 27% | [15%, 39%] | 38% | [18%, 57%] | 14% | [5%, 22%] | 13% | [3%, 24%] | 35% | [20%, 51%] |
|  | Los Angeles | 30% | [24%, 36%] | 32% | [25%, 39%] | 20% | [13%, 26%] | 11% | [7%, 15%] | 38% | [32%, 44%] |
|  | Monterey | 26% | [14%, 38%] | 27% | [15%, 38%] | 15% | [6%, 24%] | 12% | [4%, 20%] | 47% | [31%, 64%] |
|  | San Diego | 23% | [17%, 29%] | 35% | [27%, 43%] | 18% | [11%, 24%] | 5% | [3%, 7%] | 42% | [36%, 48%] |
|  | Shasta | 22% | [16%, 28%] | 36% | [26%, 46%] | 17% | [10%, 23%] | 5% | [3%, 7%] | 42% | [36%, 49%] |
| Age | 18-25 | 40% | [18%, 62%] | 33% | [13%, 52%] | 33% | [11%, 54%] | 8% | [2%, 13%] | 27% | [13%, 42%] |
|  | 26-40 | 31% | [21%, 41%] | 34% | [25%, 44%] | 22% | [13%, 31%] | 9% | [6%, 13%] | 34% | [27%, 42%] |
|  | 41-65 | 26% | [20%, 33%] | 33% | [27%, 39%] | 17% | [12%, 22%] | 9% | [5%, 14%] | 41% | [34%, 47%] |
|  | 65+ | 13% | [7%, 19%] | 28% | [17%, 40%] | 7% | [3%, 11%] | 6% | [2%, 10%] | 59% | [48%, 69%] |
| Race/Ethnicity | Latinx | 36% | [27%, 45%] | 24% | [17%, 31%] | 23% | [15%, 30%] | 13% | [8%, 18%] | 40% | [33%, 48%] |
|  | Non-Latinx (all races) | 18% | [15%, 22%] | 41% | [34%, 47%] | 14% | [10%, 17%] | 4% | [3%, 6%] | 41% | [37%, 45%] |
|  | Non-Latinx White | 12% | [12%, 20%] | 44% | [37%, 52%] | 12% | [7%, 16%] | 4% | [3%, 6%] | 40% | [35%, 45%] |
|  | Non-Latinx Asian | 21% | [12%, 30%] | 26% | [15%, 37%] | 16% | [7%, 25%] | 3% | [3%, 7%] | 53% | [44%, 61%] |
|  | Non-Latinx Black | 16% | [7%, 25%] | 53% | [27%, 79%] | 12% | [4%, 21%] | 4% | [1%, 6%] | 31% | [18%, 45%] |
| Healthy Places Index | Quartile 1 | 30% | [21%, 40%] | 29% | [19%, 38%] | 16% | [8%, 24%] | 15% | [9%, 21%] | 41% | [32%, 50%] |
|  | Quartile 2 | 28% | [20%, 37%] | 33% | [25%, 41%] | 21% | [14%, 29%] | 7% | [4%, 10%] | 39% | [32%, 45%] |
|  | Quartile 3 | 23% | [16%, 30%] | 42% | [32%, 52%] | 17% | [11%, 24%] | 6% | [3%, 8%] | 35% | [29%, 42%] |
|  | Quartile 4 | 20% | [12%, 28%] | 27% | [20%, 35%] | 17% | [9%, 25%] | 3% | [2%, 5%] | 52% | [46%, 59%] |

**TABLE 4.** CHILD SEROSTATUS BY REGION, AGE, RACE/ETHNICITY. AND HPI QUARTILE

| Characteristic |  | Evidence of Prior Infection |  | Serostatus |  |  |  |  |  |  |  |
| --- | --- | --- | --- | --- | --- | --- | --- | --- | --- | --- | --- |
|  |  | Weighted Percent | [95% CI] | Seronegative |  | Prior Infection Only |  | Prior Infection and Vaccination |  | Vaccination Only |  |
|  |  |  |  | Weighted Percent | [95% CI] | Weighted Percent | [95% CI] | Weighted Percent | [95% CI] | Weighted Percent | [95% CI] |
| Overall |  | 30% | [24%, 36%] | 57% | [48%, 66%] | 26% | [19%, 32%] | 5% | [1%, 8%] | 13% | [7%, 18%] |
| County | Alameda | 15% | [1%, 29%] | 68% | [35%, 100%] | 15% | [0%, 30%] | 0% | [0%, 0%] | 17% | [0%, 40%] |
|  | El Dorado | 19% | [7%, 31%] | 69% | [40%, 97%] | 19% | [5%, 33%] | 0% | [0%, 1%] | 12% | [6%, 19%] |
|  | Kern | 18% | [6%, 30%] | 63% | [35%, 92%] | 17% | [7%, 26%] | 2% | [0%, 4%] | 18% | [0%, 43%] |
|  | Los Angeles | 37% | [25%, 49%] | 51% | [34%, 67%] | 28% | [15%, 41%] | 9% | [0%, 18%] | 12% | [4%, 21%] |
|  | Monterey | 39% | [19%, 59%] | 56% | [27%, 85%] | 34% | [12%, 56%] | 5% | [0%, 13%] | 6% | [0%, 16%] |
|  | San Diego | 31% | [17%, 45%] | 57% | [38%, 76%] | 29% | [11%, 47%] | 1% | [0%, 4%] | 12% | [1%, 23%] |
|  | Shasta | 27% | [13%, 41%] | 64% | [40%, 87%] | 24% | [11%, 38%] | 2% | [0%, 5%] | 9% | [4%, 15%] |
| Age | 6 months - 5 years | 29% | [7%, 51%] | 71% | [39%, 100%] | 29% | [7%, 51%] | 0% | [0%, 0%] | 0% | [0%, 0%] |
|  | 5 years - 11 years | 36% | [21%, 52%] | 64% | [46%, 81%] | 36% | [21%, 52%] | 0% | [0%, 0%] | 0% | [0%, 0%] |
|  | 12 years - 17 years | 27% | [17%, 37%] | 52% | [39%, 65%] | 20% | [12%, 28%] | 8% | [2%, 14%] | 21% | [12%, 29%] |
| Race/Ethnicity | Latinx | 35% | [23%, 46%] | 51% | [38%, 65%] | 28% | [17%, 38%] | 7% | [1%, 13%] | 14% | [6%, 22%] |
|  | Non-Latinx (all races) | 24% | [15%, 33%] | 65% | [52%, 79%] | 23% | [14%, 32%] | 1% | [0%, 3%] | 10% | [5%, 16%] |
|  | Non-Latinx White | 26% | [15%, 38%] | 66% | [50%, 82%] | 26% | [14%, 37%] | 1% | [0%, 1%] | 8% | [6%, 10%] |
|  | Non-Latinx Asian | 21% | [3%, 39%] | 52% | [27%, 77%] | 20% | [2%, 38%] | 1% | [0%, 2%] | 27% | [2%, 52%] |
|  | Non-Latinx Black | 39% | [10%, 68%] | 59% | [2%, 94%] | 35% | [7%, 64%] | 3% | [0%, 10%] | 2% | [1%, 4%] |
| Healthy Places Index | Quartile 1 | 38% | [23%, 54%] | 47% | [29%, 64%] | 29% | [16%, 42%] | 9% | [1%, 17%] | 15% | [5%,26%] |
|  | Quartile 2 | 29% | [15%, 43%] | 58% | [40%, 75%] | 25% | [13%, 38%] | 4% | [0%, 10%] | 13% | [3%, 24%] |
|  | Quartile 3 | 23% | [10%, 37%] | 70% | [46%, 94%] | 23% | [9%, 37%] | 0% | [0%, 1%] | 7% | [4%, 10%] |
|  | Quartile 4 | 21% | [9%, 34%] | 68% | [46%, 89%] | 21% | [8%, 33%] | 1% | [0%, 1%] | 11% | [7%, 15%] |

Serostatus also varied across age groups, with the lowest susceptibility among people >65 years (28%, 95% CI [17%, 40%]) and highest proportion seronegative in children <5 years old (71%, 95% CI [40%, 100%]). Seropositivity due to vaccination alone was highest in people > 65 years (59%, 95% CI [48%, 69%]) whereas people between ages 18 and 25 years were more likely to have antibodies from prior infection alone (33%, 95% CI [11%, 54%]). When comparing across race and ethnicity, the lowest percent seronegative was in Latinx adults (24%, 95% CI [17%, 46%]); non-Latinx Asian adults were most likely to have antibodies due to vaccination alone (53%, 95% CI [44%, 61%]). (Table 3)

Finally, adults living in HPI quartiles 1 or 4 were less likely to be seronegative than adults living in HPI quartiles 2 or 3. (Table 3) In contrast, children living in HPI quartile 1 were less likely to be seronegative compared to those in the higher HPI quartiles. (Table 4)

### Evidence of Prior Infection and Infection-to-Case Ratio

Overall, 27% (95% CI [23%, 31%]) of adults and 30% (95% CI [24%, 36%]) of children had evidence of prior infection. In contrast, 11% of adults and 6% of children were confirmed COVID-19 cases as of May 8, 2021 in the COVID-19 case registry in the seven CalScope counties. The estimated infection-to-case ratio was 2.6 (95% CI [2.2, 2.9]) for adults and 5.0 (95% CI [4.0, 5.0]) for children. (Figure 3)

**Figure 3.**
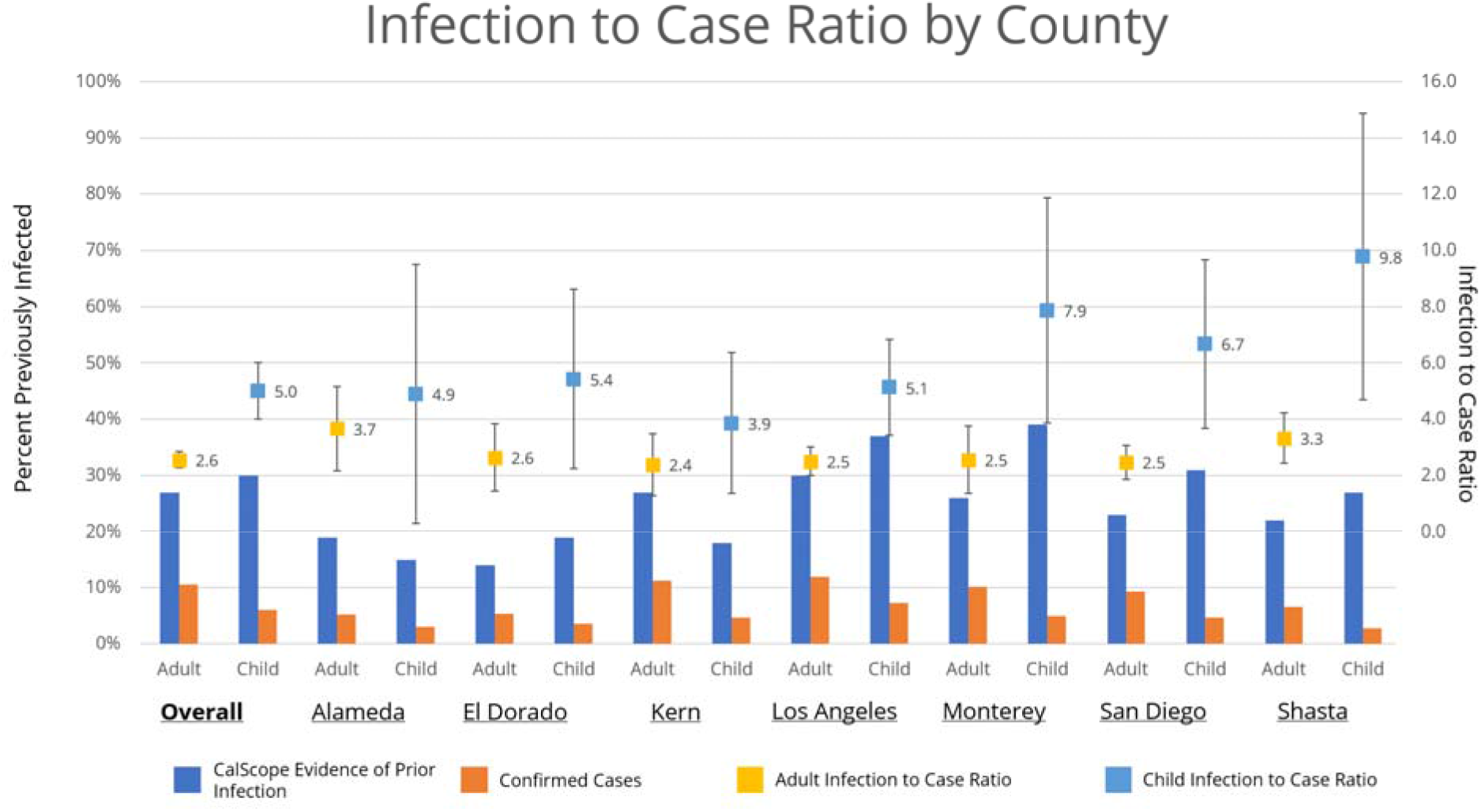
Infection to Case Ratio by County. The infection to case ratio is the ratio of the percent of the population with evidence of prior infection based on antibody test results to the percent of the population with a PCR-confirmed infection in CDPH’s COVID-19 surveillance database with an episode date on or before May 8, 2020.

Evidence of prior infection among adults varied across the seven counties from 14% (95% CI [8%, 20%]) in El Dorado County to 30% (95% CI [24%, 36%]) in Los Angeles County. Similarly, the percent of children with antibody evidence of prior infection varied from 15% (95% CI [1%, 29%]) in Alameda County to 39% (95% CI [19%, 59%]) in Monterey County. Infection-to-case ratios were consistently higher for children than adults in all counties.

Adults 18-25 years old were most likely to have evidence of prior infection (40%, 95% CI [18%, 62%]). (Table 3) Among children, those between 5 and 11 years old were most likely to have evidence of prior infection (36%, 95% CI [21%, 51%]). (Table 4) Adults >65 years were least likely to have evidence of prior infection (13%, 95% CI [7%, 19%]).

Latinx adults and children were more likely to have antibodies from prior infection (adults: 36%, 95% CI [27%, 45%]; children: 35%, 95% CI [23%, 46%]) compared with non-Latinx adults or children, and non-Latinx Asian adults and children were least likely to have antibody evidence of prior infection (adults: 21%, 95% CI [12%, 30%]; children: 21%, 95%CI [3%, 39%]).

Finally, seroprevalence of antibodies from prior infection was highest among adults and children living in the lowest HPI quartile and was lowest in adults and children living in neighborhoods in the highest HPI quartile.

## DISCUSSION

During Wave 1 of CalScope, 33% of adults and 56% of children did not have antibodies against SARS-CoV-2 as of June 2021, with 27% of adults and 30% of children having evidence of prior SARS-CoV-2 infection. Overall, the infection-to-case ratio was 2.6 for adults and 5.0 for children suggesting that through June 2021, similar numbers of infections had occurred in adults and children, but infections in children were less likely to be diagnosed because of less widespread testing in children.

Serostatus differed across region, race/ethnicity, age, and HPI quartile reflecting disparate patterns of infection and vaccination. For example, California has been prioritizing equity in its COVID-19 response by using the HPI to target vaccination campaigns, testing, and other COVID-19 mitigation measures towards more disadvantaged neighborhoods, which have borne a larger burden of the COVID-19 pandemic thus far.^29^ Our results suggest that these targeted vaccination campaigns have been effective – seropositivity due to vaccination is similar for adults in the lowest and highest HPI quartiles (56% in both).

To the best of our knowledge, this is the first study to conduct population-based serological testing of children under 18 in the United States. Though the proportion of children and adults who had been previously infected was similar, most children remained seronegative as of June 2021 because they were not yet eligible for vaccination. Even among those eligible (age 12-17), vaccination coverage has been low, and many were still seronegative. With in-person schooling resuming in much of the state, it will be important to encourage vaccination of all age eligible children.

The ADAP assays used in CalScope are highly sensitive and specific, but the assays have only been validated up to 4 months post-infection. Thus, we do not know the extent of antibody waning below the limit of detection after >4 months. This means that our estimates of seroprevalence due to prior infection might underestimate true cumulative incidence—particularly in populations who were infected in the Spring of 2020. Additionally, individuals whose antibodies wane below detectable levels after vaccination or infection may not be equally susceptible to subsequent infection or COVID-19 disease as immunologically naïve individuals because of cell-mediated immunity.^30^ So, our seroprevalence estimates may underestimate the proportion of the population with immunity.

We anticipated that those who enrolled in our study might not be representative of our target populations, so we used causal transportability to design our study and survey instrument to generalize our results to our target populations. Our weighted results can be considered unbiased estimates of SARS-CoV-2 serostatus in our target population if we assume that we were able to measure and adjust for all the differences between the study sample and target population that were associated with SARS-CoV-2 serostatus. If our weights excluded any key characteristics that differed between the sample and target population and that affected SARS-CoV-2 serostatus, our results might still suffer from residual non-response bias. However, our estimates are in line with those from other studies and known patterns of vaccination and infection, so residual biases are unlikely to meaningfully affect our results.

Overall, we found that similar proportions of adults and children had been infected through June 2021, but serostatus varied substantially across region, age group, and by race/ethnicity. Though seroprevalence studies such as CalScope are unable to measure all aspects of the immune response, spike antibodies are a correlate of protection for SARS-CoV-2 infection and symptomatic disease.^31–33^ As vaccination and transmission continues, the population that remains most vulnerable to COVID-19 infection and disease will evolve. It is critical that public health agencies monitor who does not have SARS-CoV-2 antibodies to accurately forecast future COVID-19 surges. CalScope will begin collecting data for Wave 2 in Fall 2021 and Wave 3 will occur in the first half of 2022.

## Data Availability

All data produced in the present study are available upon reasonable request to the authors

## ACKNOWLEDGEMENTS

We thank Daniela Valenzuela, Evelyn Cubias, Mauricio Ollervides, Edgar Martinez, Kara Gionfriddo, Lourdes Mariana Ponte-Cordova, and Stefanie Medlin for their work in addressing participant questions and concerns. We thank Thomas Quarre and Siddarth Satish for their support in designing and maintaining the CalScope website. The study was funded by a CDC ELC grant to the California Department of Public Health.

---

**SUPPLEMENTARY FIGURE 1.**
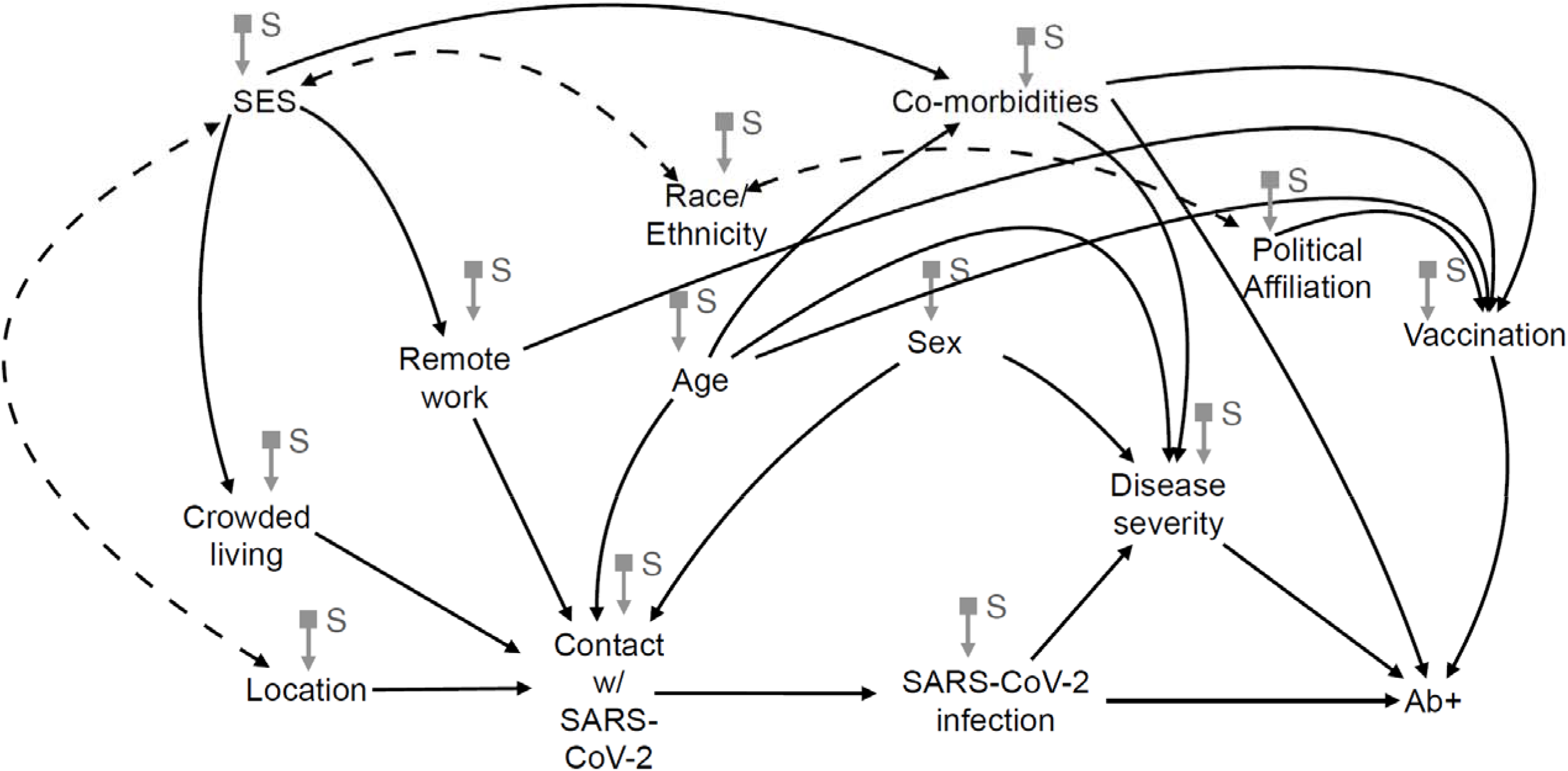

**SUPPLEMENTARY FIGURE 2.**
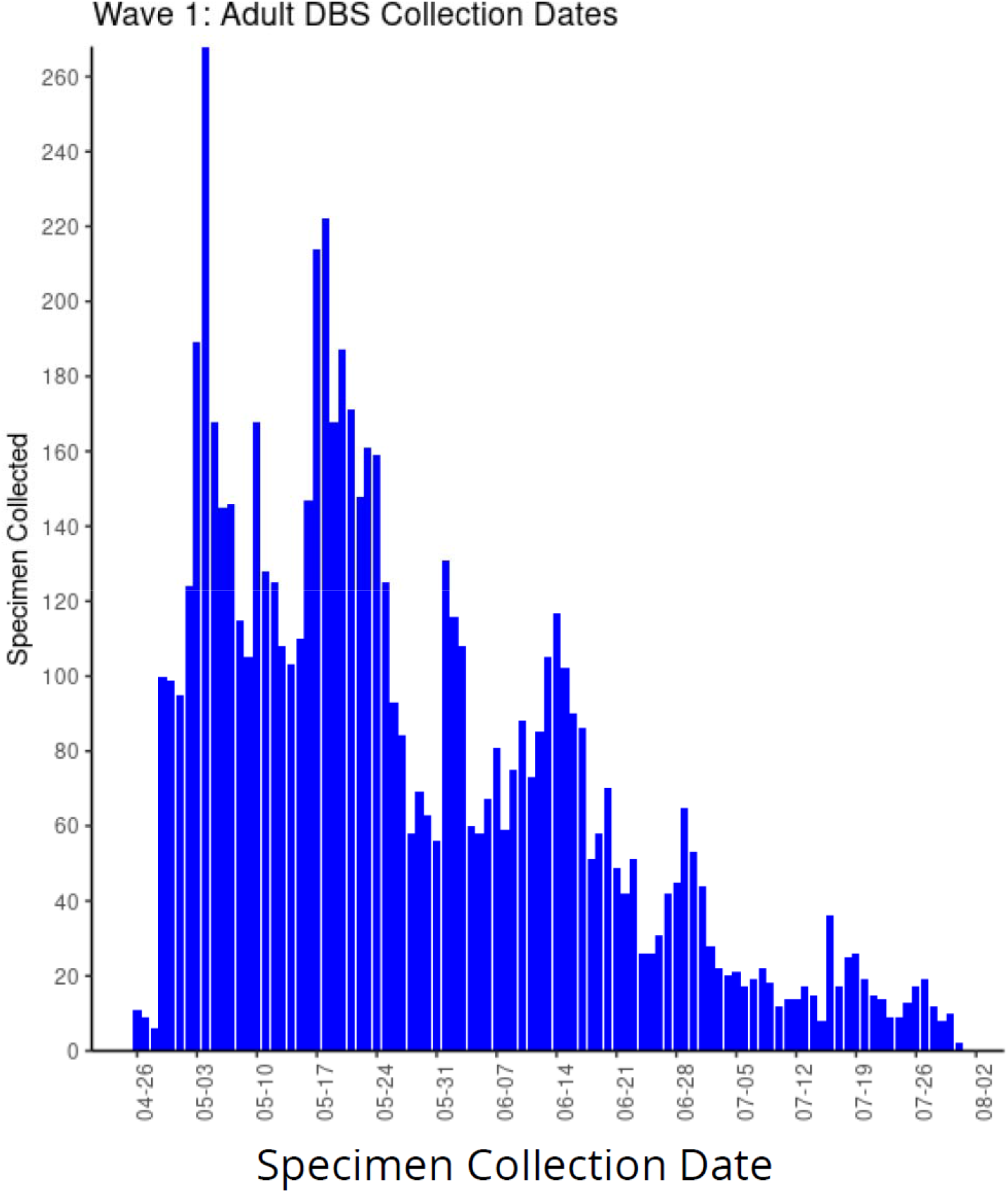

## APPENDIX

### COUNTY SELECTION

First, we divided the state into 7 mutually exclusive geographic regions such that the counties within each region were similar with respect to SARS-CoV-2 cumulative confirmed cases and known risk factors for SARS-CoV-2. Cumulative confirmed cases were based on PCR-confirmed COVID-19 cases in the California’s COVID-19 surveillance database as of November 2020. Census-tract level measures hypothesized to be associated with prior and ongoing SARS-CoV-2 infection risk included poverty and crowded living conditions derived from the 2015 American Community Survey and the University of California San Francisco Health Atlas^1, 2^

Next, one county was selected to represent each region. To ensure that estimates from each county could be generalized to their respective regions, we selected the county that was most heterogeneous with respect to SARS-CoV-2 risk factors, race, ethnicity, age, and cumulative confirmed COVID-19 cases relative to the other counties within the region.^3^

### ASSAY PROCEDURES

Complete details of the ADAP assay have been published previously.^4^ The DBS specimen were eluted by punching six 3mm discs into 1mL of elution buffer and incubating at 37°C for 90min. The eluents were then concentrated with 100kD MWCO filter column at 14,000 rcf for 9min. The eluents were then mixed with spike and nucleocapsid protein-DNA conjugates, followed with enzymatic ligation to reunite nearby DNA within the antibody-antigen immune complex. The ligated products were then amplified with PCR and quantified with specific primers in 384-well qPCR.

## Adult-Child Survey

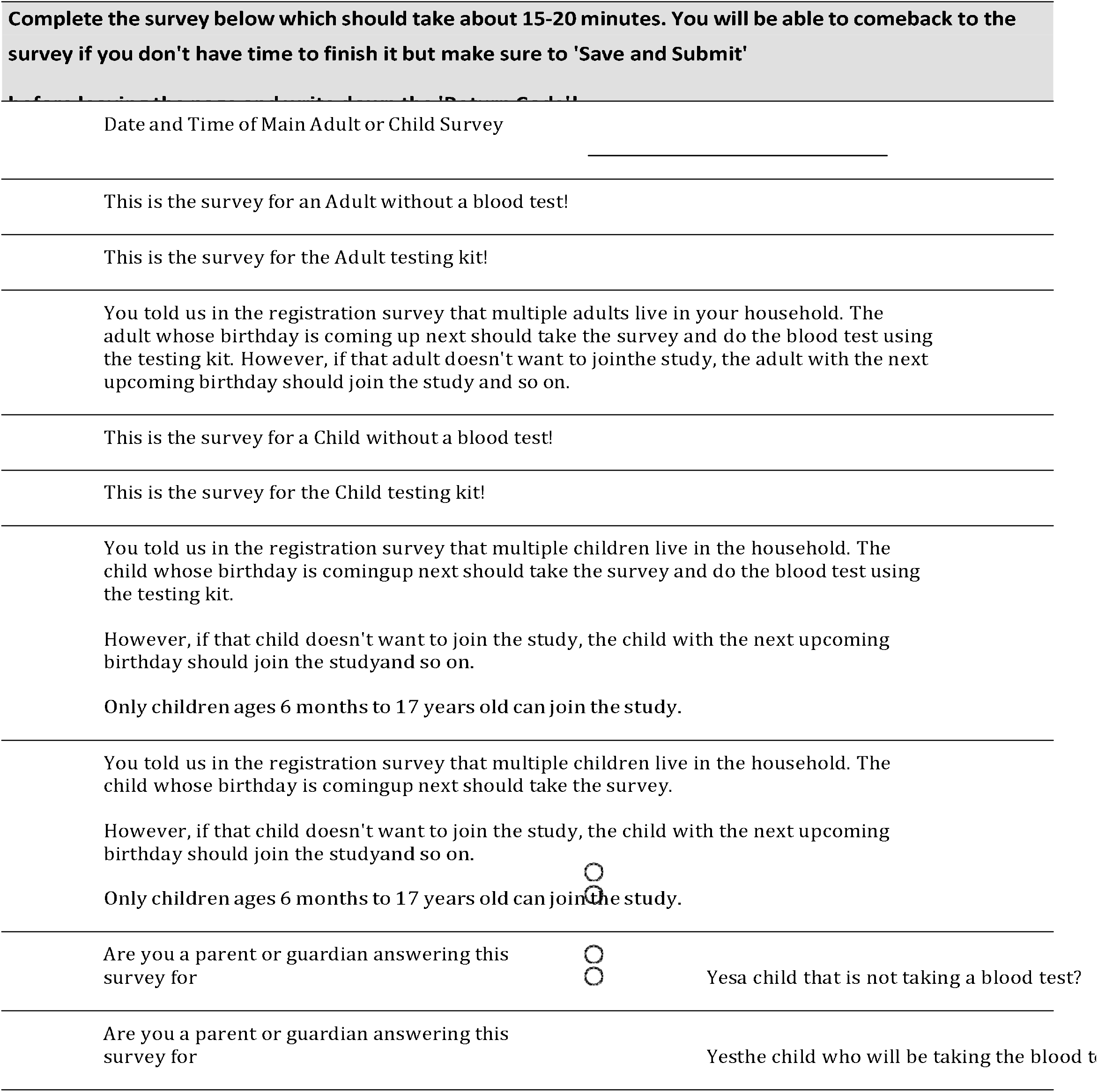
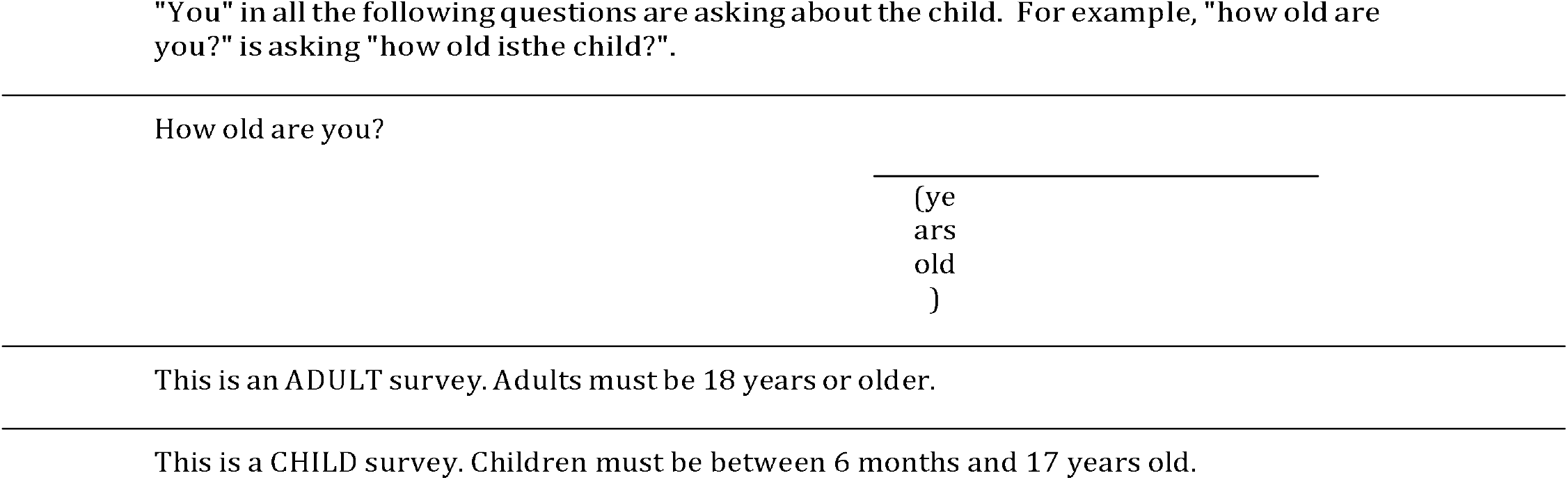
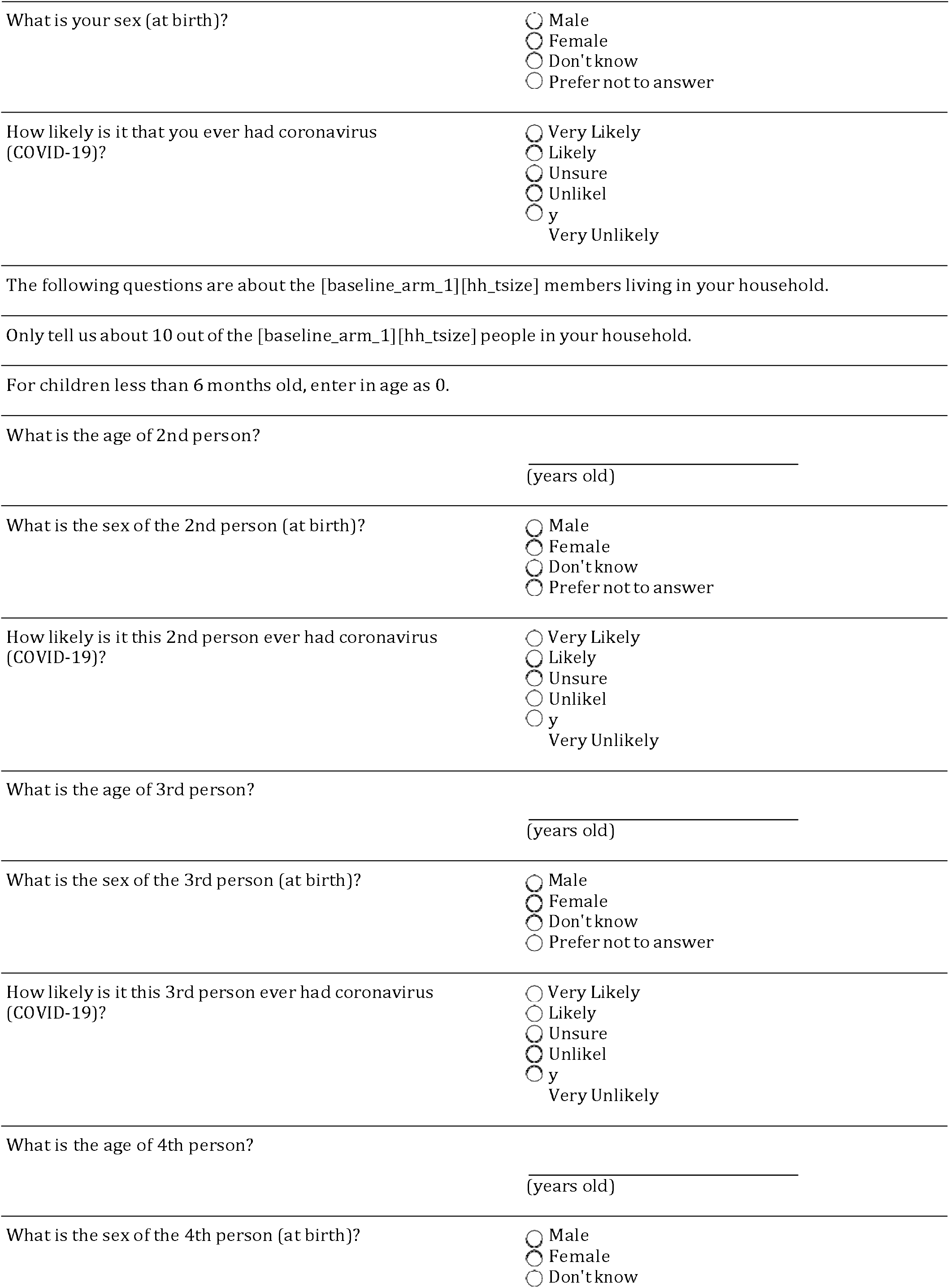
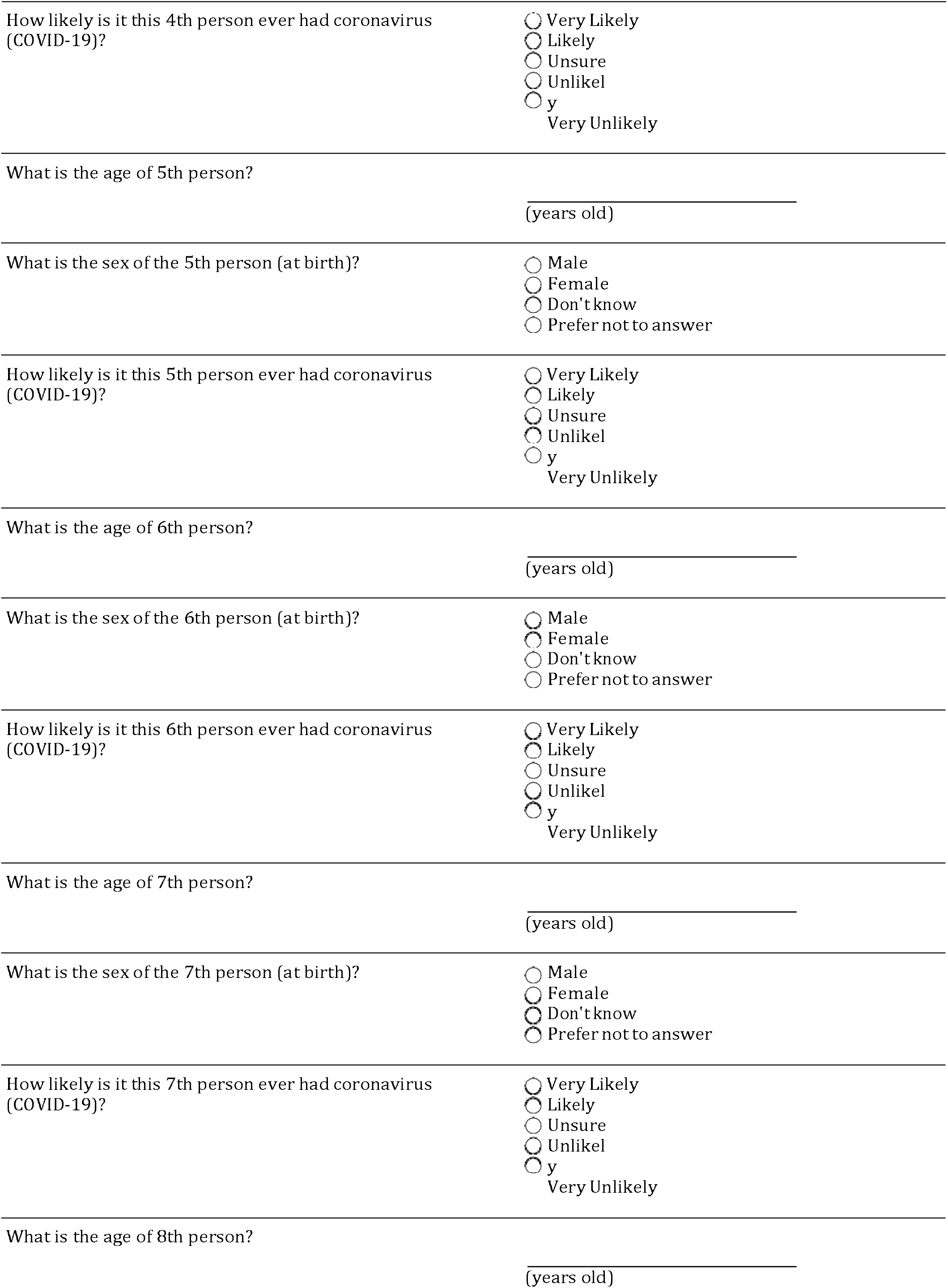
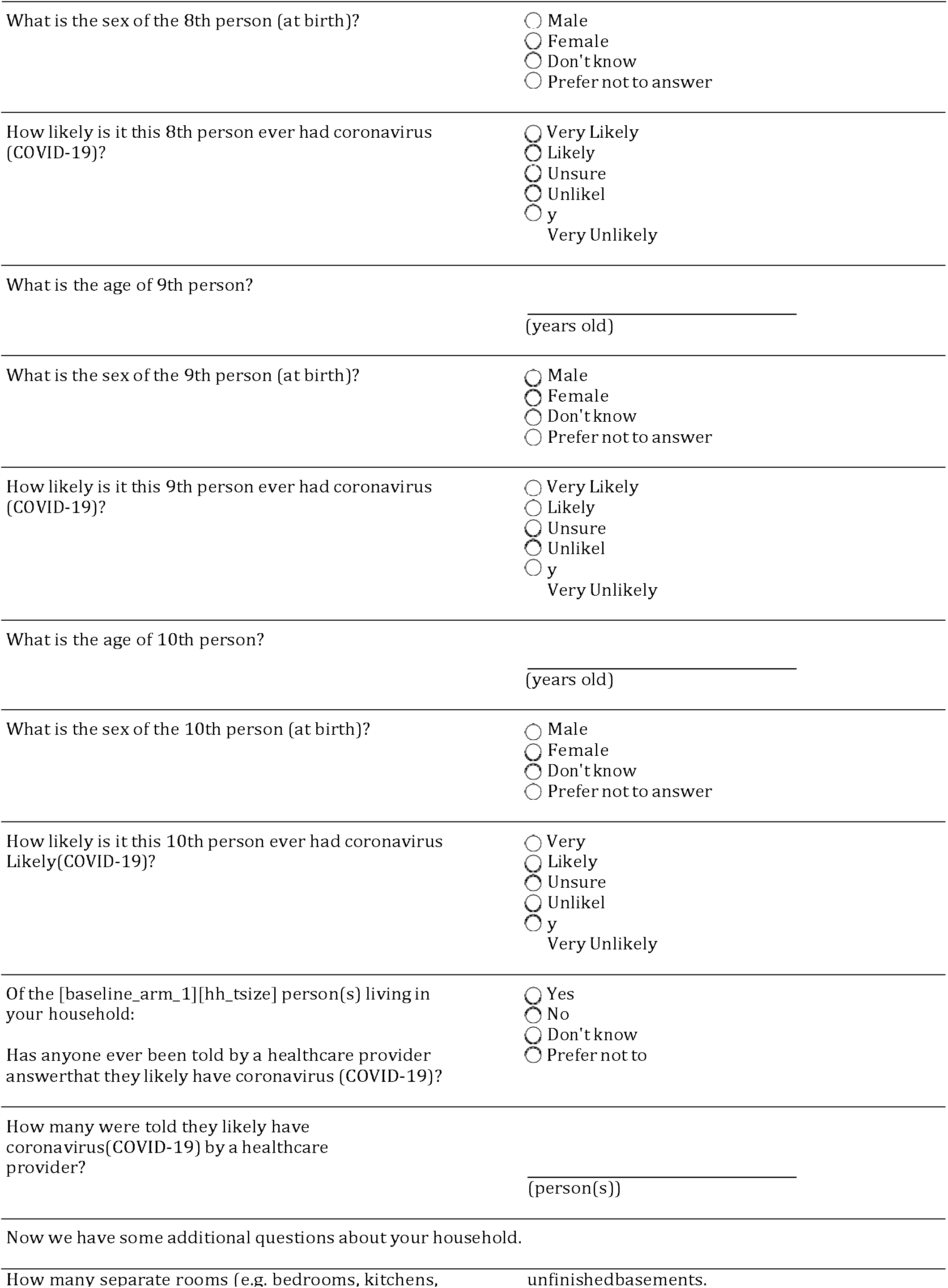
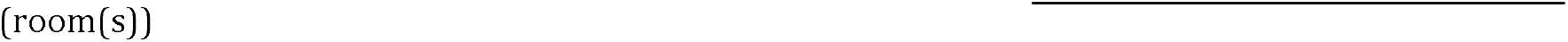
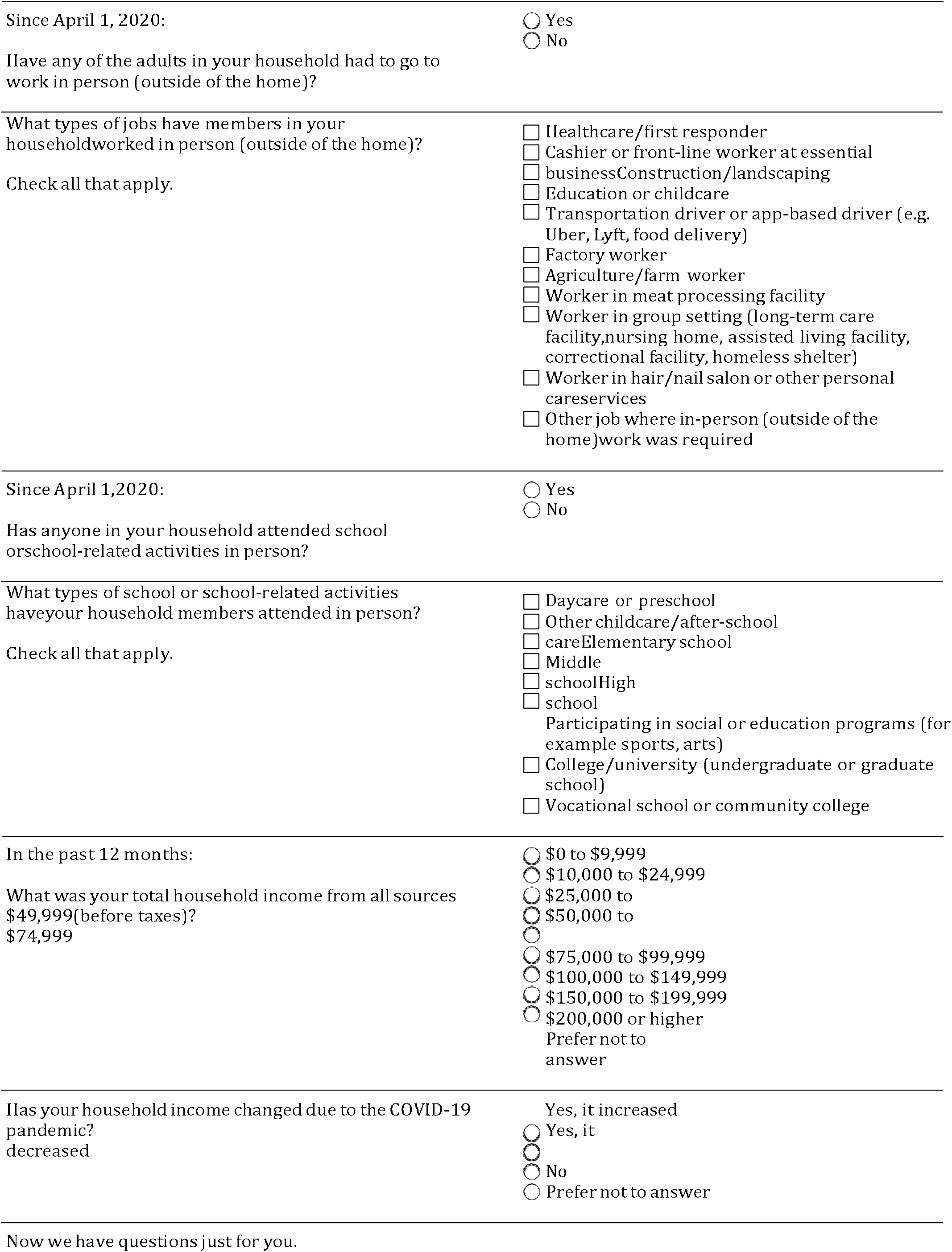
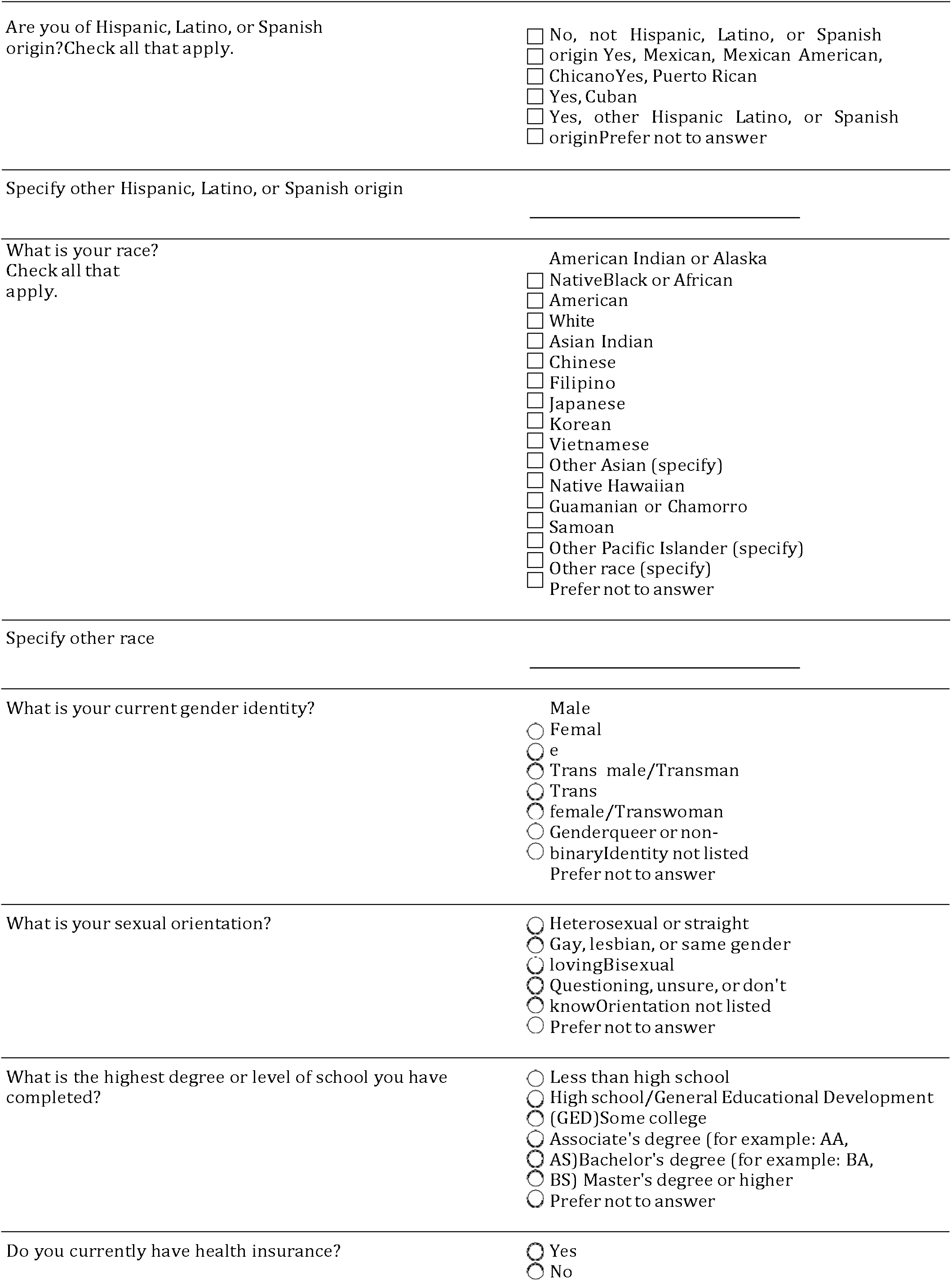
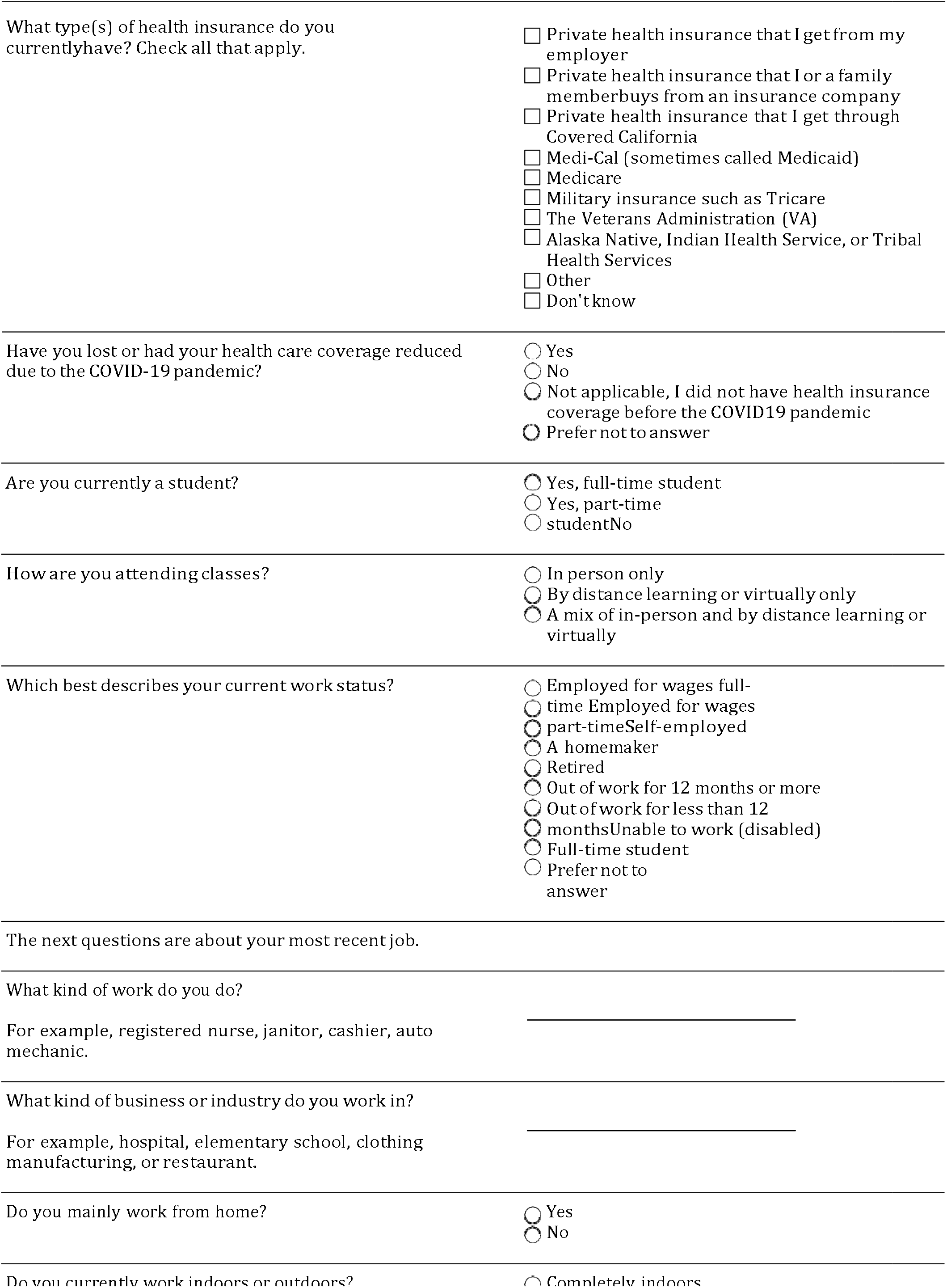
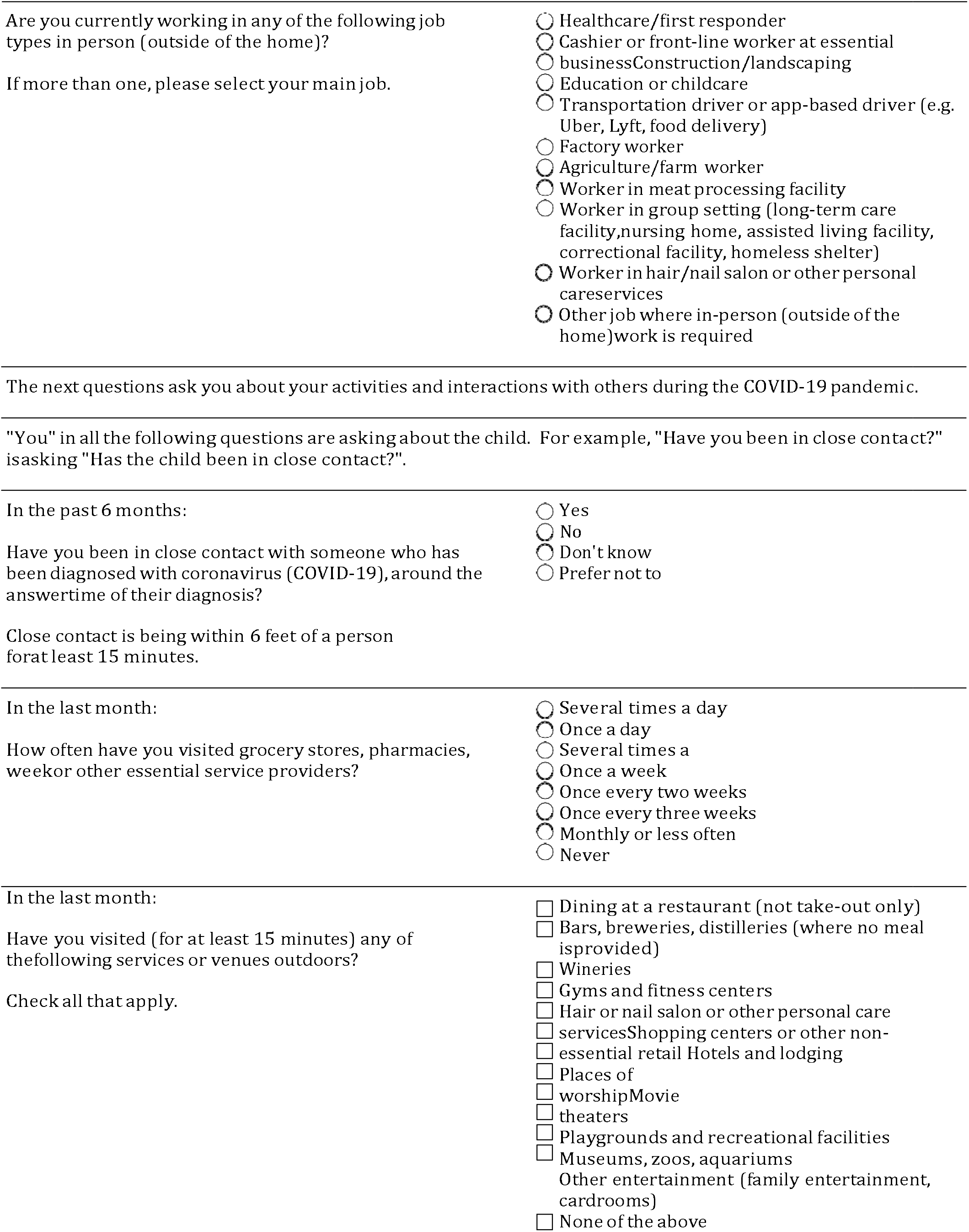
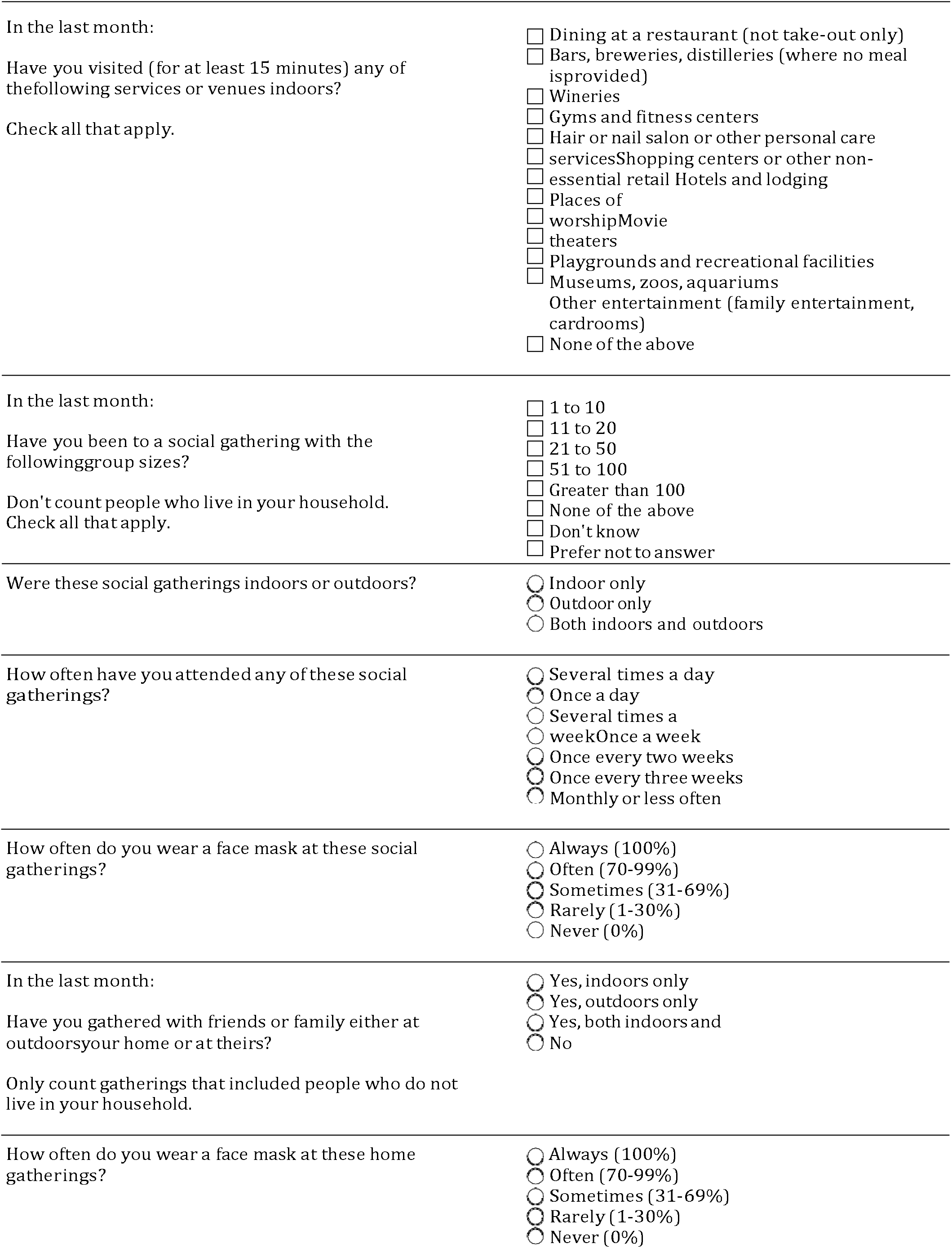
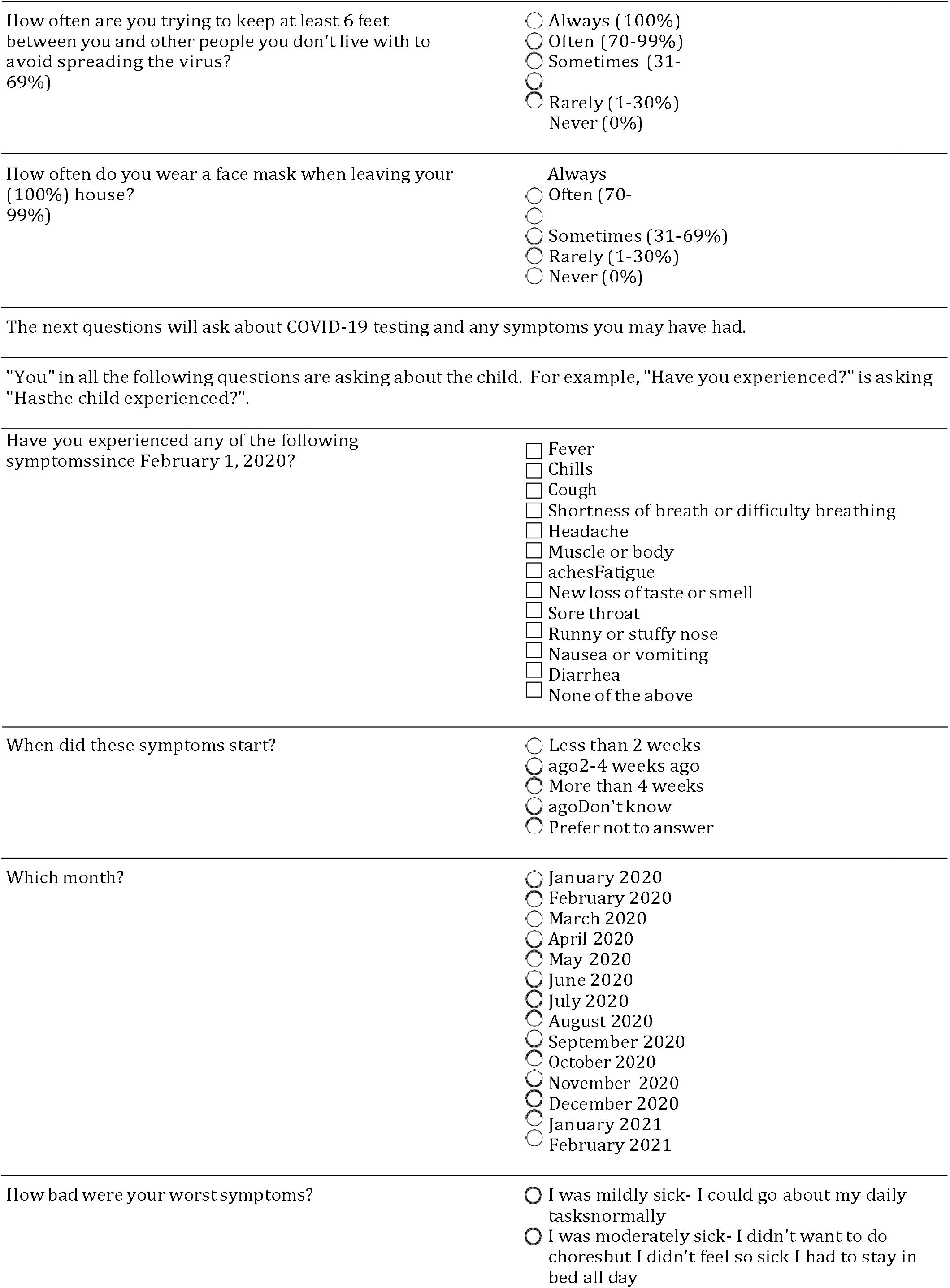
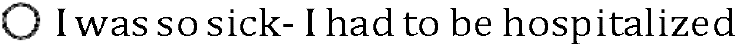
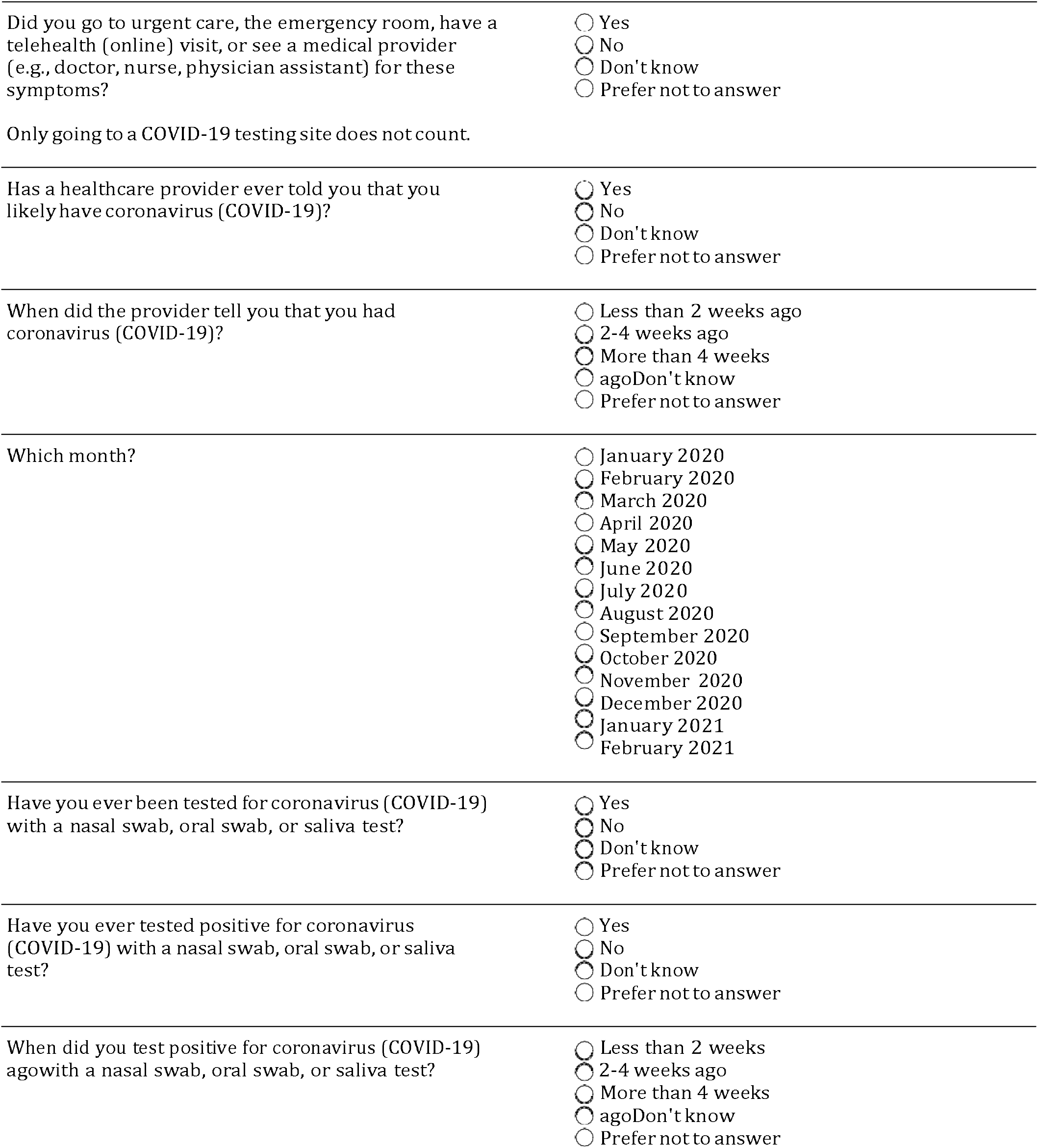
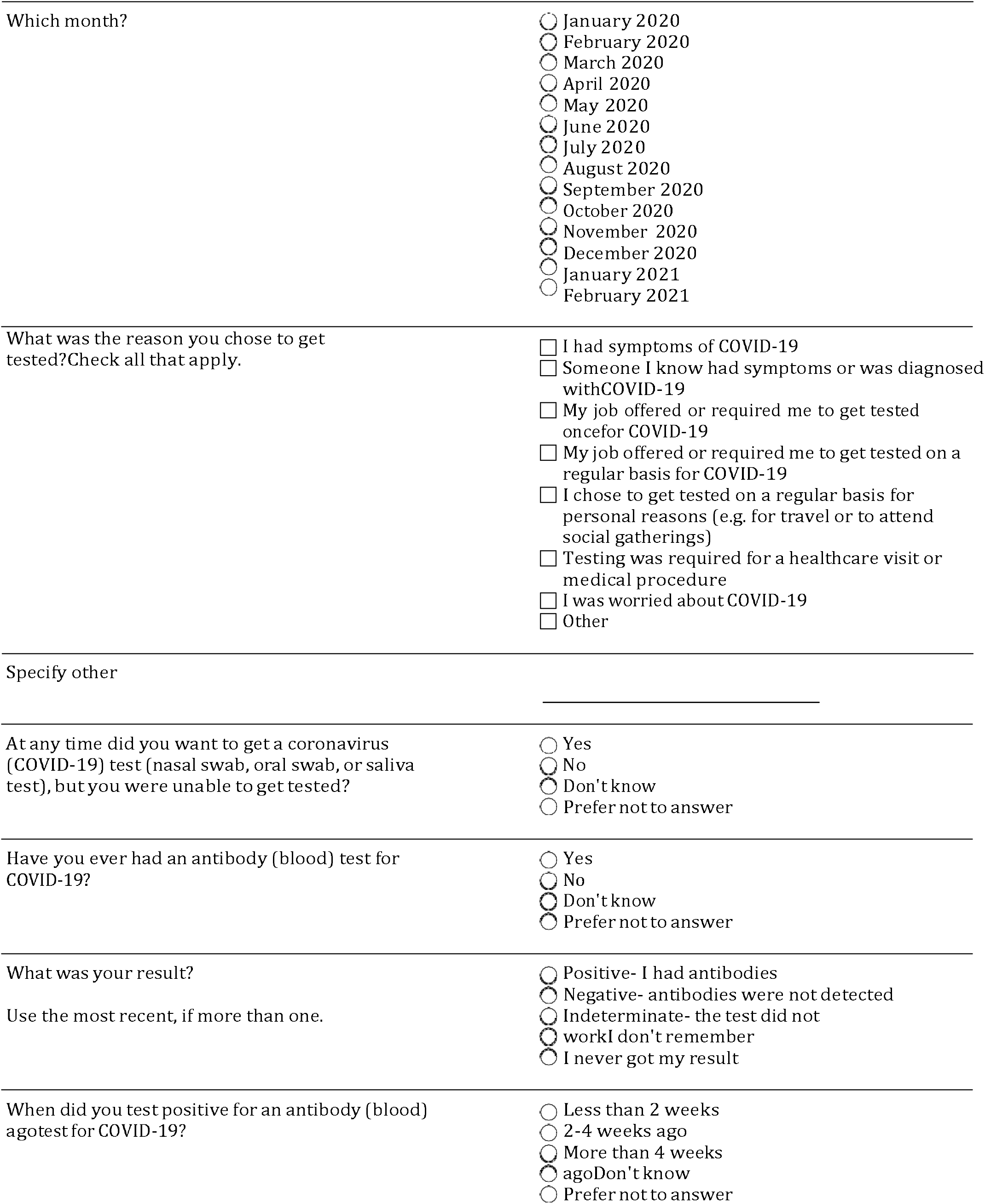
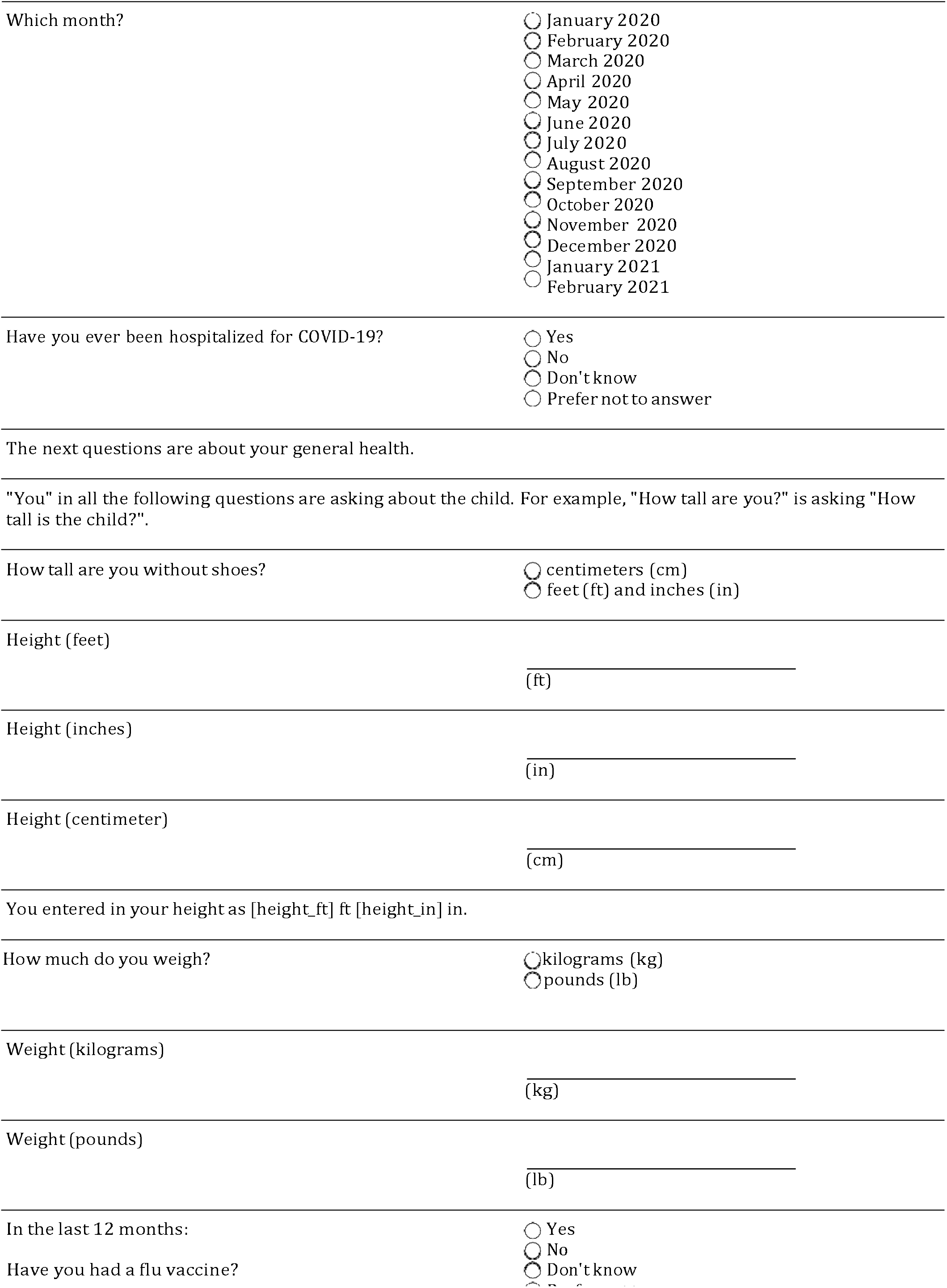
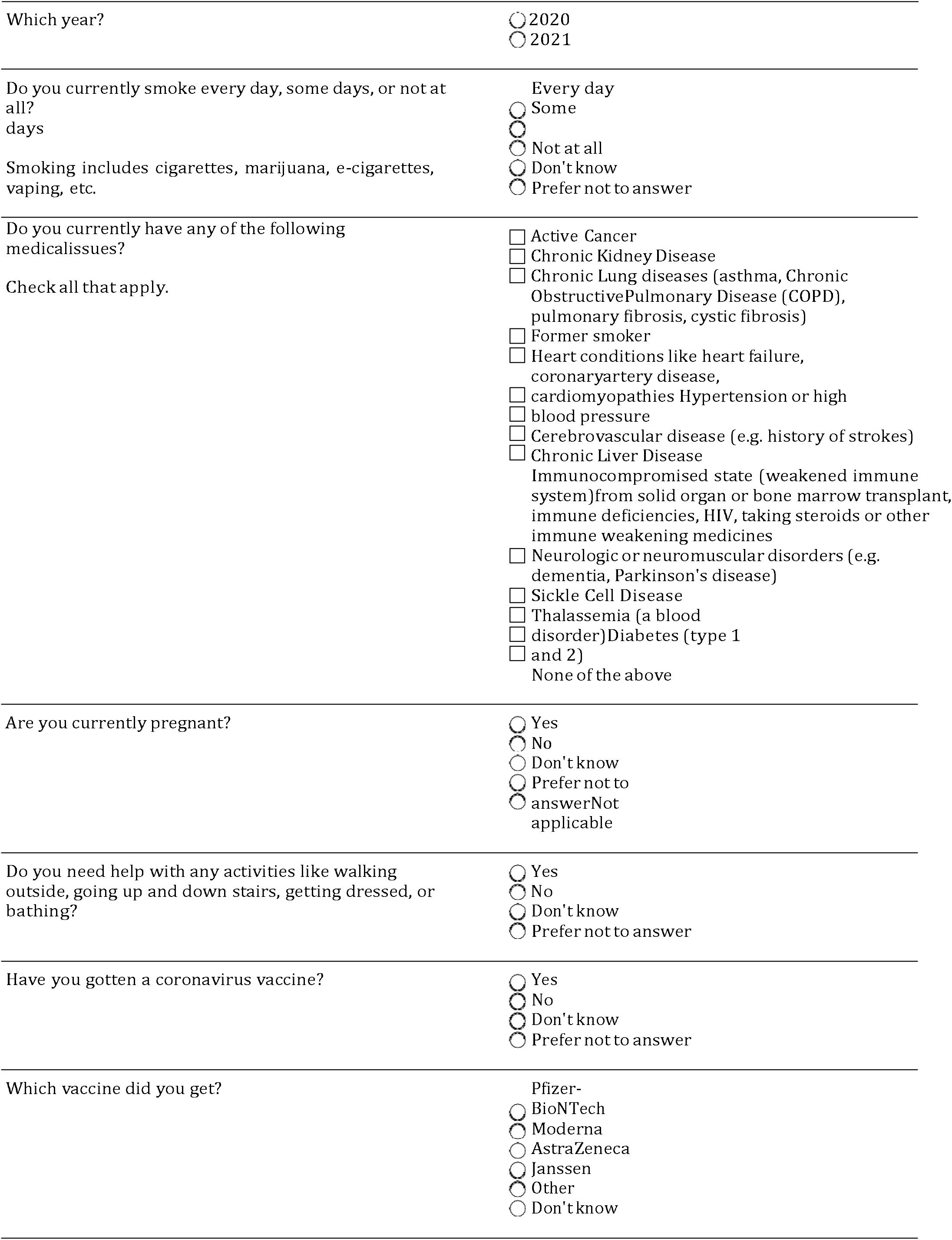
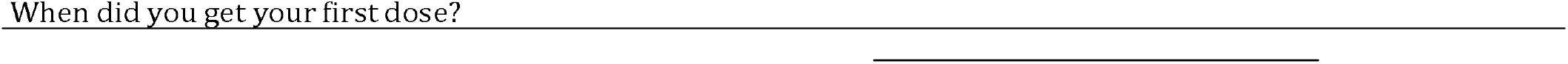
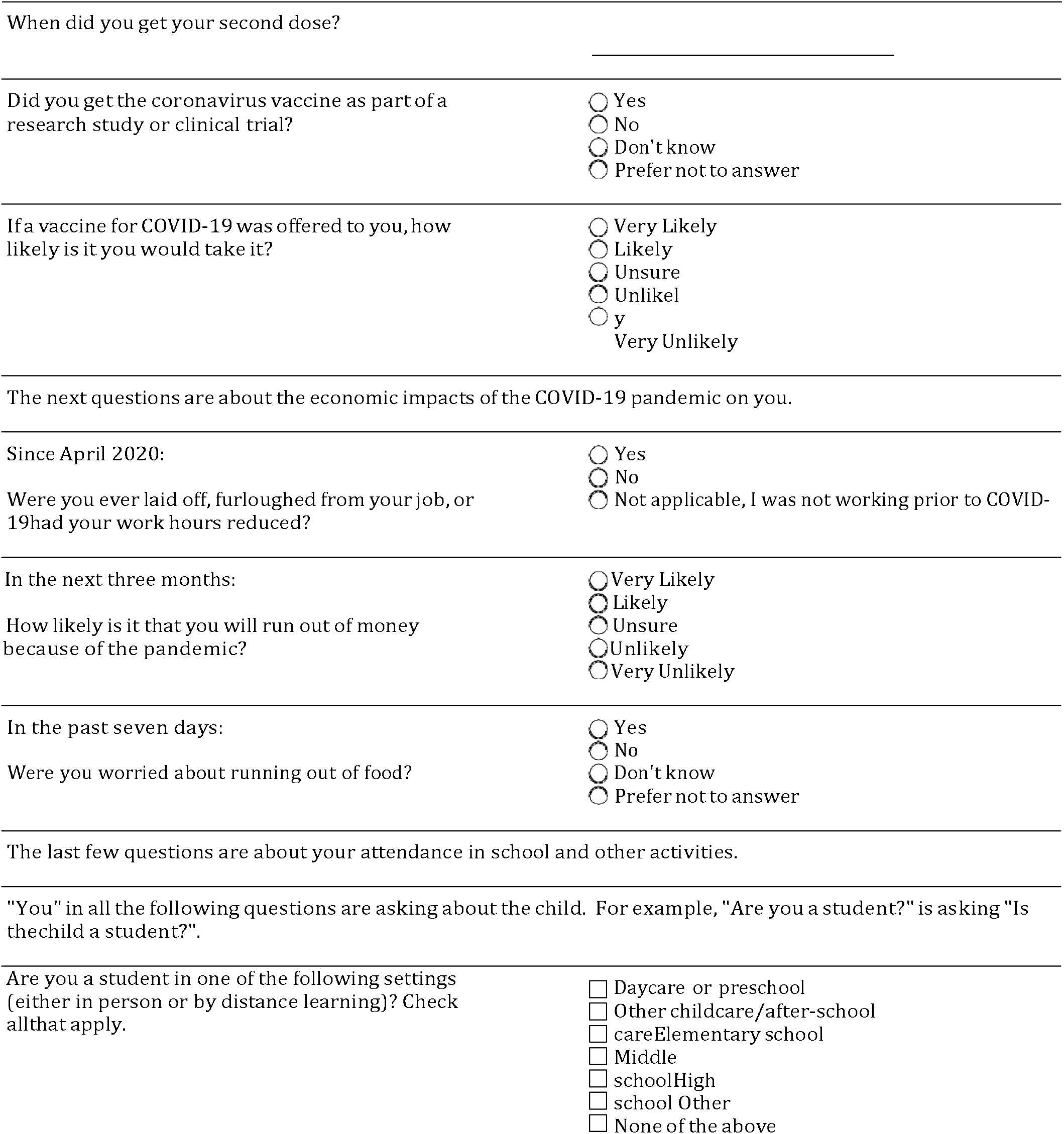
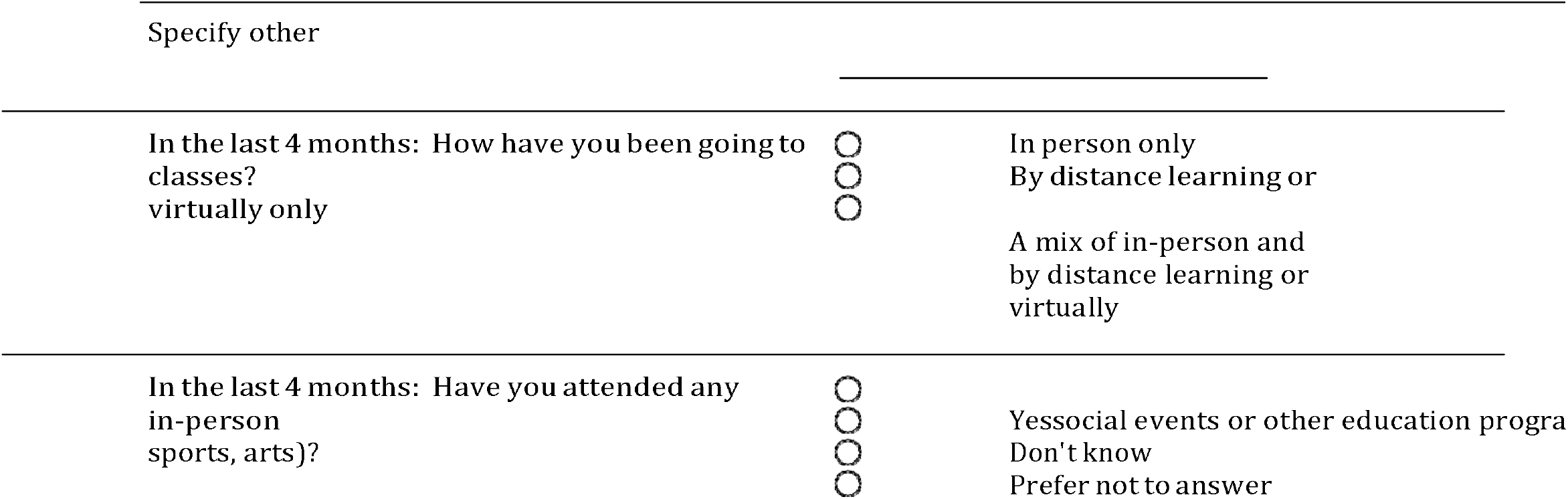

## REFERENCES

1. United States COVID-19 Cases and Deaths by State over Time | Data | Centers for Disease Control and Prevention. https://data.cdc.gov/Case-Surveillance/United-States-COVID-19-Cases-and-Deaths-by-State-o/9mfq-cb36 (accessed Oct 4, 2021).

2. CalCAT. https://calcat.covid19.ca.gov/cacovidmodels/ (accessed Oct 4, 2021).

3. Baden LR, El Sahly HM, Essink B, et al. Efficacy and Safety of the mRNA-1273 SARS-CoV-2 Vaccine. New England Journal of Medicine 2021; 384: 403–16.

4. Polack FP, Thomas SJ, Kitchin N, et al. Safety and Efficacy of the BNT162b2 mRNA Covid-19 Vaccine. New England Journal of Medicine 2020; 383: 2603–15.

5. Gazit S, Shlezinger R, Perez G, et al. Comparing SARS-CoV-2 natural immunity to vaccine-induced immunity: reinfections versus breakthrough infections. Infectious Diseases (except HIV/AIDS), 2021 DOI:10.1101/2021.08.24.21262415.

6. Lamba K, Bradley H, Shioda K, et al. SARS-CoV-2 Cumulative Incidence and Period Seroprevalence: Results From a Statewide Population-Based Serosurvey in California. Open Forum Infectious Diseases 2021; 8. DOI:10.1093/ofid/ofab379.

7. Robertson MM, Kulkarni SG, Rane M, et al. Cohort profile: a national, community-based prospective cohort study of SARS-CoV-2 pandemic outcomes in the USA—the CHASING COVID Cohort study. BMJ Open 2021; 11: e048778.

8. Siegler AJ, Sullivan PS, Sanchez T, et al. Protocol for a national probability survey using home specimen collection methods to assess prevalence and incidence of SARS-CoV-2 infection and antibody response. Annals of Epidemiology 2020; 49: 50–60.

9. Sullivan PS, Siegler AJ, Shioda K, et al. Severe Acute Respiratory Syndrome Coronavirus 2 Cumulative Incidence, United States, August 2020–December 2020. Clinical Infectious Diseases 2021; published online July 10. DOI:10.1093/cid/ciab626.

10. Bajema KL, Wiegand RE, Cuffe K, et al. Estimated SARS-CoV-2 Seroprevalence in the US as of September 2020. JAMA Intern Med 2020; published online Nov 24. DOI:10.1001/jamainternmed.2020.7976.

11. Havers FP, Reed C, Lim T, et al. Seroprevalence of Antibodies to SARS-CoV-2 in 10 Sites in the United States, March 23-May 12, 2020. JAMA Intern Med 2020; published online July 21. DOI:10.1001/jamainternmed.2020.4130.

12. Ng DL, Goldgof GM, Shy BR, et al. SARS-CoV-2 seroprevalence and neutralizing activity in donor and patient blood. Nature Communications 2020; 11: 4698.

13. Anand S, Montez-Rath M, Han J, et al. Prevalence of SARS-CoV-2 antibodies in a large nationwide sample of patients on dialysis in the USA: a cross-sectional study. Lancet 2020; 396: 1335–44.

14. Bendavid E, Mulaney B, Sood N, et al. COVID-19 antibody seroprevalence in Santa Clara County, California. International Journal of Epidemiology 2021; 50: 410–9.

15. Sood N, Simon P, Ebner P, et al. Seroprevalence of SARS-CoV-2–Specific Antibodies Among Adults in Los Angeles County, California, on April 10-11, 2020. JAMA 2020; 323: 2425–7.

16. Pearl J, Bareinboim E. Transportability of Causal and Statistical Relations: A Formal Approach. In: 2011 IEEE 11th International Conference on Data Mining Workshops. Vancouver, BC, Canada: IEEE, 2011: 540–7.

17. U.S. Census Bureau. American Community Survey (2015). 2015.

18. Karp DG, Danh K, Espinoza NF, Seftel D, Robinson PV, Tsai C. A serological assay to detect SARS-CoV-2 antibodies in at-home collected finger-prick dried blood spots. Sci Rep 2020; 10: 20188.

19. Pearl J, Bareinboim E. External Validity: From Do-Calculus to Transportability Across Populations. Statistical Science 2014; 29: 579–95.

20. Cole SR, Stuart EA. Generalizing Evidence From Randomized Clinical Trials to Target Populations: The ACTG 320 Trial. American Journal of Epidemiology (England) 2010; 172: 107–15.

21. Petersen ML. Compound Treatments, Transportability, and the Structural Causal Model: The Power and Simplicity of Causal Graphs. Epidemiology (Cambridge, Mass) 2011; 22: 378–81.

22. California S of. Essential workforce. https://covid19.ca.gov/essential-workforce/ (accessed Oct 6, 2021).

23. Statewide Database. 2020 General Election Precinct Data. www.statewidedatabase.org/d10/g20.html (accessed Aug 5, 2021).

24. Public Health Alliance of Southern California. The California Healthy Places Index (HPI). 2021; published online April 22. www.healthyplaces.org (accessed Aug 5, 2021).

25. van der Laan MJ, Rose S. Targeted Learning. New York, NY: Springer New York, 2011 http://link.springer.com/10.1007/978-1-4419-9782-1 (accessed Oct 23, 2016).

26. Bacharach M. Estimating Nonnegative Matrices from Marginal Data. International Economic Review 1965; 6: 294–310.

27. Pipelines for Machine Learning and Super Learning. https://tlverse.org/sl3/index.html (accessed Oct 5, 2021).

28. R Core Team. R: A Language and Environment for Statistical Computing. Vienna, Austria: R Foundation for Statistical Computing, 2018 https://www.R-project.org/.

29. Blueprint for a Safer Economy. https://www.cdph.ca.gov/Programs/CID/DCDC/Pages/COVID-19/COVID19CountyMonitoringOverview.aspx (accessed Oct 5, 2021).

30. Cassaniti I, Percivalle E, Bergami F, et al. SARS-CoV-2 specific T-cell immunity in COVID-19 convalescent patients and unexposed controls measured by ex vivo ELISpot assay. Clinical Microbiology and Infection 2021; 27: 1029–34.

31. Corbett KS, Nason MC, Flach B, et al. Immune correlates of protection by mRNA-1273 vaccine against SARS-CoV-2 in nonhuman primates. Science; 373: eabj0299.

32. Harvey RA, Rassen JA, Kabelac CA, et al. Association of SARS-CoV-2 Seropositive Antibody Test With Risk of Future Infection. JAMA Internal Medicine 2021; 181: 672–9.

33. Addetia A, Crawford KHD, Dingens A, et al. Neutralizing Antibodies Correlate with Protection from SARS-CoV-2 in Humans during a Fishery Vessel Outbreak with a High Attack Rate. J Clin Microbiol 2020; 58: e02107–20.

## APPENDIX REFERENCES

1. Emeruwa UN, Ona S, Shaman JL, et al. Associations Between Built Environment, Neighborhood Socioeconomic Status, and SARS-CoV-2 Infection Among Pregnant Women in New York City. JAMA. 2020;324(4):390. doi:10.1001/jama.2020.11370

2. UCSF Health Atlas. Accessed October 7, 2020. https://healthatlas.ucsf.edu

3. Rudolph KE, Díaz I, Rosenblum M, Stuart EA. Estimating Population Treatment Effects From a Survey Subsample. Am J Epidemiol. Published online September 4, 2014:kwu197. doi:10.1093/aje/kwu197

4. Karp DG, Danh K, Espinoza NF, Seftel D, Robinson PV, Tsai C ting. A serological assay to detect SARS-CoV-2 antibodies in at-home collected finger-prick dried blood spots. Sci Rep. 2020;10(1):20188. doi:10.1038/s41598-020-76913-6

